# The Impact of Gender on Pediatric Surgical Access and Outcomes in Africa

**DOI:** 10.1101/2024.02.10.24302627

**Authors:** Sacha Williams, Olivia Serhan, Jenny Wang, Christian Guindi, Elena Guadagno, Maeve Trudeau, Emannuel Ameh, Kokila Lakhoo, Dan Poenaru

## Abstract

**Introduction:** Girls, whose care is often affected by barriers steeped in gender inequity, may be at higher risk of poor surgical outcomes. This study explored the impact of gender on pediatric surgical care in Africa.

**Methods:** Differences in access to care and clinical outcomes for boys and girls were examined for pediatric surgical conditions that do not differ by physiological sex. A systematic review of African pediatric surgical studies ensued, followed by a random effects meta-analysis, and risk of bias assessment.

**Results:** Of the 12281 records retrieved, 54 were selected for review. Most studies were retrospective (57.4%), single-site (94.4%), from Egypt, Nigeria, Ghana, or Ethiopia (55.6%), focussed on gastrointestinal conditions (63.0%), published in 2010 or sooner (85.1%), had study durations of 5 years or less (68.5%), and cohorts of less than 200 children (57.4%). Sixty percent reported the outcome of mortality. Meta-analysis odds ratios revealed surgery was performed 3.6 times more often on boys (95% CI: 2.6, 4.9); and mortality was 1.6 times greater for girls (95% CI: 1.3, 2.0).

**Conclusion:** African girls appear to face gender inequities in pediatric surgical care. Findings will be further explored in a mixed-methods study.

**Level of evidence:** **I**

**Highlights:**

- Gender disparities in global surgical care have been documented in the African adult population. However gender specific differentials in surgical access and outcomes have yet to be documented for African pediatric populations.
- This study provides first-time evidence of gender inequity in pediatric surgical care in Africa.

## Introduction

Children remain among the most vulnerable populations worldwide. As their numbers rapidly expand, the global burden of disease also increases [1–9]. In fact, estimates suggest that currently 1.7 billion children lack access to surgery that is both safe and affordable [10]. A significant proportion of these children reside in low- and middle-income countries (LMICs), that are frequently operating with limited resources and infrastructure [11,12]. In Africa, the population demographic is approximately 50% pediatric; by corollary, the surgical disease burden is critical [11,12]. However, the surgical care of children is an aspect of child health that continues to be inadequate, despite its potential for reducing or even eliminating child morbidity and/or mortality [13].

*Gender* is a social construct that may further compound the issue. It can be defined as a set of characteristics, roles, and behaviors ascribed to perceptions of masculinity and femininity [13,14], potentially mutable according to times and/or society norms and values. While gender may intersect with *sex*, they are not necessarily the same, as the latter tends to focus on biological factors like chromosomes, hormones, or reproductive organs. [14,15] Historically, gender has been implicated in hierarchical power imbalances, whereby, within the male-female dichotomy, females were deemed inferior [15]. As a result, gender norms may influence children’s health and well-being. For example, in certain African societies, gender inequity has been reported to manifest as social and/or structural barriers negatively impacting females [15]. Moreover, recent reports have suggested that, in some African countries, women seek care for their surgical conditions less commonly than men, and those who do undergo surgery experience poorer surgical outcomes [16–20].

Despite the evidence of gender disparity in the adult African population, potential gender differentials in the surgical care of African children have not been documented to date. This study sought to address the knowledge gap by exploring whether there were gender disparities in access, and postoperative outcomes for African children requiring surgery.

## Methods

### Caveats, Definitions

Although gender and biological sex may overlap, they are not necessarily the same, as sex is centered solely upon biological factors (e.g. reproductive organs, chromosomes etc). However, since global databases, registries, and studies continue to classify cohorts according to sex instead of gender, sex was used as a proxy for gender, particularly in cases where *conditions should not have sex differences, since they manifest equally in boys and girls* (equal sex ratio). There would be no biological, physiological, nor anatomical reason for differences in incidence by sex. Any conditions specific to boys (e.g. male circumcision, testicular torsion, etc.), or girls alone (e.g. ovarian cysts, pregnancy, etc.) were excluded from the analysis in this study. Only gender neutral pediatric surgical conditions were examined. Therefore, any deviations from standard 1:1 or predicted sex ratios in the conditions studied were attributed to gender-based factors. This methodology was confirmed with leading chairs in sex and gender research, an Equity Diversity and Inclusion (EDI) Advisor, and our African collaborators. Cis- or transmales or females were not evaluated, as these distinctions were not reported in the included studies.

The study explored the primary outcomes of reported access to care and postoperative outcomes. Access was defined as the number of actual surgical interventions performed by sex. Surgical outcomes included the following: mortality, surgical site infection (SSI), and any other reported adverse effects or complications.

### Registration and Reporting Checklist

The study protocol was registered with PROSPERO (CRD42023395021), the international registry for systematic reviews [21], and conducted in accordance with the guidelines of the Preferred Reporting Items for Systematic Reviews and Meta-Analyses (PRISMA) statement [22].

### Search Strategy and Information Sources

A comprehensive electronic literature search was conducted by a senior medical librarian using the following databases from inception through December 2022 with no language restrictions: Medline (Ovid), Embase (Ovid), CINAHL (EBSCO), Cochrane (Wiley), Global Health (Ovid), Web of Science (Clarivate Analytics), Africa Wide Information (EBSCO) and Global Index Medicus (WHO). The search strategy involved text words from the title, abstract, keywords, or subject headings to retrieve papers on gender inequality, pediatric surgery, and surgical care in Africa, without language restriction. Various gender and sex terms were searched, including sex/gender characteristics or factors, gender identity, and minority groups. Conference abstracts, books and book chapters were excluded in Embase. The full search strategy is located in **Appendix A.** PRISMA and PRISMA-S extension were used and are located in **Appendix B**

### Inclusion and Exclusion Criteria

Study inclusion criteria were: 1. quantitative research papers, with 2. study cohorts under 19 years of age, 3. situated in an African setting, with 4. sex/gender data, surgical access and postoperative outcomes data, 5. for surgical conditions that occur equally in boys and girls. Exclusion criteria included: qualitative research, editorials/commentaries, conference abstracts, books, book chapters, populations over 19 years of age, gender-specific surgical conditions, studies outside of Africa, without sex/gender data, surgical access and/or outcomes data.

### Study Selection

Duplications of eligible studies were removed in EndNote X9 (EndNote X9 2023, Clarivate, Philadelphia, PA, USA) and uploaded into Rayyan software (Rayyan 2022, Boston, MA, USA), for literature screening by multiple independent reviewers (SW, JW, CG, OS). Titles and abstracts were first screened for meeting study eligibility. In instances of disagreement, discussion and full consensus was sought amongst reviewers. If consensus could not be found, a senior reviewer (DP) was available for adjudication. Full papers of all abstracts deemed potential for inclusion were subsequently retrieved and independently screened (SW, OS, JW) for adherence to study criteria.

### Data Collection

The PRISMA flow diagram was drafted from the finalized list of relevant pediatric surgical papers. (**Figure 1**) Pertinent data variables were extracted from the studies selected for inclusion and placed in an Excel spreadsheet (Excel 2021, Microsoft, Richmond, VA, USA). Variables included: title, authors, year, journal, country, surgical subspecialty (pediatric surgery, neurosurgery, plastic surgery, ophthalmology, urology, orthopedics, burn, trauma surgery), diagnosis/condition, procedure/intervention, duration of study, population type (pediatric, mixed), total population, mean/median age, number of males/females, male/female postoperative complications, mortality).

**Fig. 1.** PRISMA flowchart identifying eligible studies Page MJ, McKenzie JE, Bossuyt PM, Boutron I, Hoffmann TC, Mulrow CD, et al. The PRISMA 2020 statement: an
updated guideline for reporting systematic reviews. BMJ 2021;372:n71. doi: 10.1136/bmj.n71

Sex ratios (male:female) for: surgical access, postoperative complications (excluding death), mortality, and proportions of deaths by sex were calculated when not provided directly in the studies (**Appendix C**), in support of the Meta-Analysis. The rationale for the proportions of deaths by sex calculations was that the number of boys in the studies generally exceeded the number of girls. So it would enable one to see the mortality rates within each sub-cohort (e.g. male, female), in light of their vastly different total numbers.

### Meta-Analysis

Given the wide array of surgical disciplines and measured indices in the included studies, a random effects meta-analysis was conducted. Statistical analyses were performed in Jamovi v.2.3 (Jamovi Project, Sydney, Australia) and STATA 18 (StataCorp, 2023, College Station, TX, USA) to ensure accuracy, and for graphical representations [23,24]. Results were calculated in Excel (Excel 2021, Microsoft, Richmond, VA, USA) to confirm accuracy. An alpha level of p < 0.05 indicated statistical significance. The log odds ratio was utilized as the outcome measure. Heterogeneity (Tau^2^) was approximated via the restricted maximum-likelihood estimator [24]. The I^2^, H^2^ statistic, and Cochrane’s Q test were also calculated to assess heterogeneity. Prediction intervals for true outcomes were calculated for I^2^ > 40% and/or tau^2^ > 0 as applicable. Forest plots were created for variables, access and postoperative outcomes to display significance. Analysis in the form of studentized residuals and Cook’s distances were performed to identify outliers and/or influential studies in the model. Those with a studentized residual greater than the 100 x (1 – 0.05/2k)th percentile of a normal distribution were considered to be potential outliers. Studies were deemed influential if they had a Cook’s distance greater than the median plus six times the interquartile range of the Cook’s distances. Funnel plots were created to assess publication bias. Potential funnel plot asymmetry was verified via the rank correlation, and regression tests.

### Quality Assessment

The quality of the evidence of the studies (prospective, cross-sectional, case review) was analyzed using the Risk of Bias in Non-Randomized Studies – of Interventions (ROBINS-I) tool [25]. This was done to evaluate risk of bias in estimates of safety or efficacy of interventions employed in non-randomized studies. The ROBINS-I instrument evaluated domains pre- and at-intervention (bias due to confounding, in the selection of participants into the study, and in the classification of outcomes), post-intervention (bias due to deviations from intended interventions and missing data, in outcomes measurement, and in the selection of reported results). Decisions were made on a scale ranging from: low, moderate, serious, critical risk of bias, or no information.

## Results

### Study Selection

The PRISMA flowchart is illustrated in **Figure 1**. The electronic literature search in the eight databases yielded 12,281 records. After duplicates were removed using EndNote, 10,579 records were available for screening. Titles/abstracts were screened in Rayyan for adherence to study criteria and resulted in 1,534 abstracts selected for potential inclusion. Two papers could not be retrieved, resulting in 1,532 eligible reports. Retrieval of full papers and subsequent review yielded 54 sex-disaggregated papers selected for inclusion. Reasons for paper exclusion ranged from: sole focus on access or outcomes (n = 460, n = 2 respectively), obstetrics and gynecology cases (n = 337), female genital mutilation (n = 265), male sexual health (n = 241), female-specific fistula (n = 83), ineligible population (n = 64), and case reports (n = 26). The finalized list of included pediatric surgical studies, involving gender neutral conditions, and their recorded variables are detailed in **Appendix D**.

### Study Characteristics

The included studies were predominantly single-center (n = 54; 94.4%), retrospective cohorts (n = 31; 57.4%), from either Egypt, Nigeria, Ghana, or Ethiopia (n = 30; 55.6%), focusing on pediatric surgery for gender neutral conditions (n = 34; 63.0%), published on or after 2010 (n = 46; 85.1%; range: 1980-2023), with a study duration less than 5 years (n = 37; 68.5%; range: < 1 year – 22 years), and with cohorts under 200 children (n = 31; 57.4%). Thirty-three (61.1%) of the papers reported the surgical outcome of mortality, while 21 studies (38.9%) reported other complications. (**Table 1**).

**Table 1.**
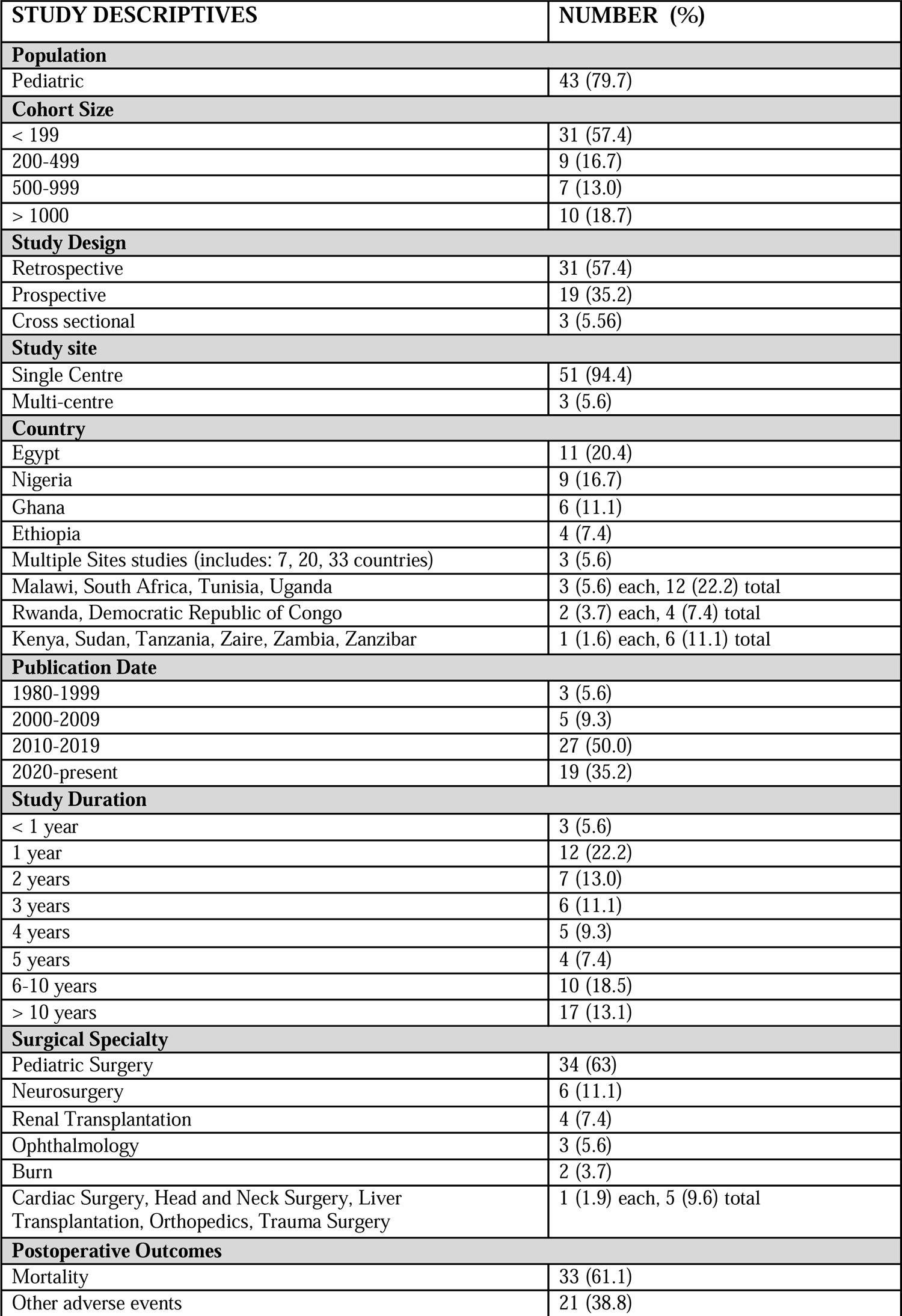
Characteristics of included studies (n = 54)

### Sex Ratios

The sex ratio (male:female) for surgical access, in these studies of gender neutral conditions, exceeded one in 90% of the cohorts (mean = 2.6), implying surgeries were predominantly performed on males. A third of the sex ratios for non-mortality postoperative complications also exceeded one, suggesting a lower rate of complications for males than females. Sixty percent of the mortality sex ratios were greater than 1 (mean = 1.9), suggesting a higher mortality rate for males than females. However, a more in depth analysis of the sex ratio for the proportions of death by sex, revealed that the majority of these values (60.0%) were below 1 (mean = 0.8). This suggested that there was a higher proportion of death among females than males. The proportion of girls that died was actually 1.3 times that of the proportion of boys, despite the expectation of gender neutrality.

### Meta-Analysis

#### A) Access

For the 54 studies included in the analysis, involving conditions that should have had the same values for boys and girls, males were more likely (94.0%) to have access to surgery than females (**Figure 2a**). Based on the random effects model, the average ratio was calculated, and found to be significantly different from zero (p < 0.0001), suggesting the results were not due to random chance. It revealed an odds ratio of 3.6 (95% CI: 2.6, 4.9), suggesting that males received surgery 3.6 times more frequently than females. Further testing indicated that there was high heterogeneity among the studies (Tau^2^ = 1.2, I^2^ = 98.7%, H^2^ = 76.5, Q = 1174.5; p < 0.001). As a result, a 95% prediction interval, or range was calculated (−0.9 – 3.5). According to studentized residuals, one study may be an outlier [24*]; and in concert with another study [15*] may be overly influential. Exploration of publication bias via funnel plot did not indicate asymmetry (**Figure 2b**). This was reinforced via Egger’s regression (0.6, p = 0.6), Spearman rank correlation (0.1, p = 0.5), and fail-safe N testing (41154, p < 0.001).

**Fig. 2.**
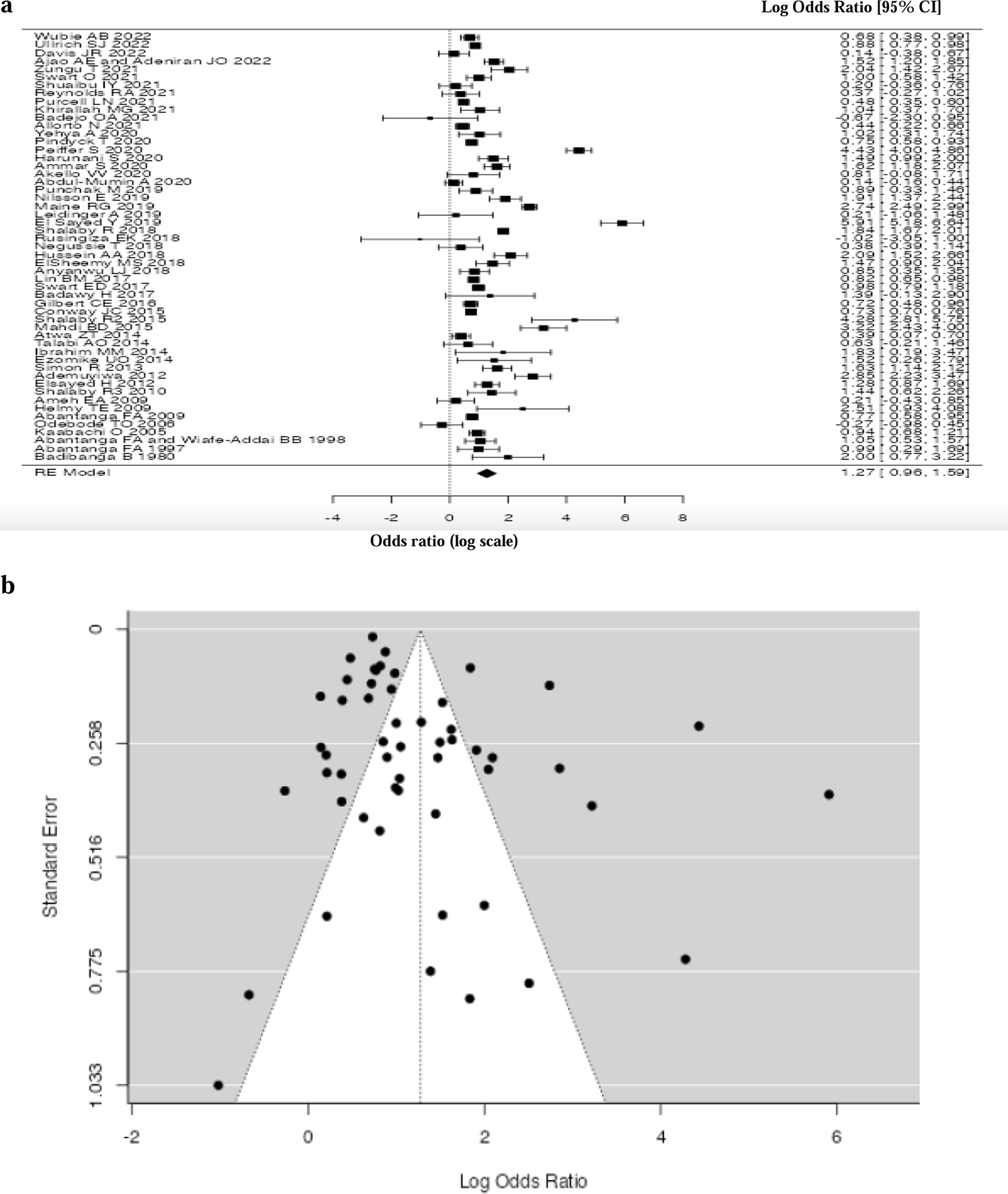
**(a)** Forest plot of access (log scale) odds ratio (n = 54); **(b)** Funnel plot representing publication bias

#### B) Adverse Postoperative Outcomes - Mortality

For the 29 studies included in the analysis, involving conditions that should have had the same values for boys and girls, mortality was higher for females than males. (76.0%). (**Figure 3a**). Based on the random effects model, the average ratio was calculated, and found to be significantly different from zero (p < 0.0001), suggesting the results were not due to random chance. It revealed an odds ratio of 1.6 (95% CI: 1.3, 2.0), suggesting that females died 1.6 times more frequently than males. Further testing indicated that there was moderate heterogeneity among the studies (Tau^2^ = 0.1, I^2^ = 41.7%, H^2^ = 1.7, Q = 41.2; p = 0.05). As a result, a 95% prediction interval, or range was calculated (−0.1 – 1.1). According to studentized residuals, there were no outliers; while another study [15*] may be overly influential [45*]. Exploration of publication bias via funnel plot did not indicate asymmetry (**Figure 3b**). This was reinforced via Egger’s regression (−0.2, p = 0.8), Spearman rank correlation (0.1, p = 0.7), and fail-safe N testing (336, p < 0.001).

**Fig. 3.**
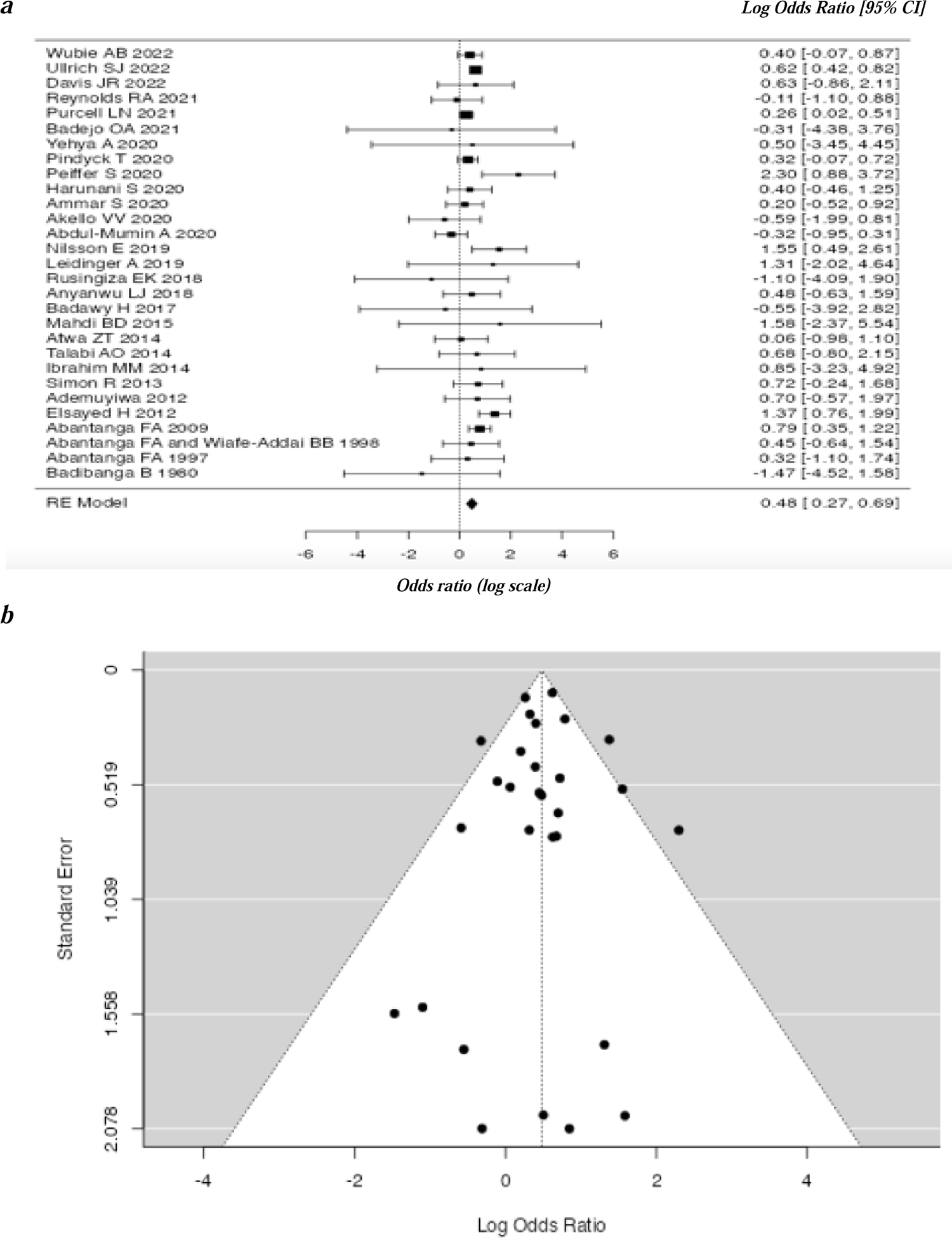
(a) Forest plot of access (log scale) odds ratio (n = 29); (b) Funnel plot representing publication bias

### Risk of Bias Assessment

Each of the 22 eligible studies for ROBINS-I assessment demonstrated a risk of bias in at least one domain (**Table 2**). Six of the 7 domains had majority judgements of low risk of bias. The domain involving bias due to confounding was the only one demonstrating critical and serious risk of biases. Across the 2 grouped domains (pre- / at-intervention, post-intervention domains), most of the bias was low to moderate (72.7-95.5% for each domain), with the former grouping (pre- / at-intervention) also containing 6 studies at serious risk of bias. One study had missing data for some domains [53*].

**Table 2.** ROBINS-I Risk of bias assessment results (n = 22)

|  | No Information | Low Risk (%) | Moderate Risk (%) | Serious Risk (%) | Critical Risk (%) |
| --- | --- | --- | --- | --- | --- |
| **Domains** |  |  |  |  |  |
| **Pre- and At-intervention** |  |  |  |  |  |
| **Bias due to confounding** | 1 (4.6) | 5 (22.7) | 4 (18.2) | 7 (31.8) | 5 (22.7) |
| **Bias in selection of participants into the study** |  | 21 (95.5) |  | 1 (4.6) |  |
| **Bias in classification of outcomes** | 1 (4.6) | 18 (81.8) | 1 (4.6) | 2 (9.1) |  |
| **Post-intervention** |  |  |  |  |  |
| **Bias due to deviations from intended interventions** | 1 (4.6) | 19 (86.36) | 2 (9.1) |  |  |
| **Bias due to missing data** | 1 (4.6) | 17 (77.3) |  | 4 (18.2) |  |
| **Bias in measurement of outcomes** |  | 21 (95.5) |  | 1 (4.6) |  |
| **Bias in selection of reported result** | 1 (4.55) | 17 (77.3) | 3 (13.6) | 1 (4.6) |  |
| **Total across pre- and at-intervention domain** |  | 8 (36.4) | 8 (36.4) | 6 (27.3) |  |
| **Total across post-intervention domain** | 1 (4.55) | 15 (68.2) | 6 (27.3) |  |  |

## Discussion

Surgery is an indispensable component of healthcare, and essential to address the burden of disease worldwide [9,26]. For every child to be granted the right to safe, timely, and affordable care, access to surgery and good postoperative outcomes are crucial. The current study has for the first time explored, via systematic review, gender disparities in surgical access and outcomes for African children. This exploration was predicated on the suspicion that gender inequities may adversely affect the health and well-being of girls.

Gender disparities have been documented to affect children in many areas, including education. As a result, the United Nations established its Sustainable Development Goals (SDGs) #4 and #5 (equitable quality education, gender equality and empowerment) to promote universal access to, and completion of, primary education, thus closing the gender gap [27]. Notwithstanding, many African countries have yet to achieve educational parity by gender [28]. According to UNESCO, one in four girls are likely to be out of school in sub-Saharan Africa [28]. In addition, these girls are more likely than their male counterparts to never enroll in school [28]. Friedman et al., in their modeling and forecast of educational inequality as it pertains to the 2030 SDG targets, reported that the education of females is predicted to lag that of males in sub-Saharan Africa through 2030 [29]. Oruko et al., in their exploration of the educational access and outcomes of Kenyan children, reported that despite the government providing free primary education, fewer girls reached secondary school level; and those that remained in school were less likely than boys to achieve a grade of C+ or better in their national exams [30]. These examples demonstrate the disadvantaged position of girls compared to boys in the educational realm.

Gender disparities in the surgical care of children have been documented tangentially in a few isolated reports. For example, Sakita et al., in their exploration of clinical profiles and outcomes of children presenting to the emergency department of a Tanzanian hospital with undifferentiated abdominal pain, reported a low proportion of girls (32.6%) in their study population [26]. Moreover, girls were associated with higher in-hospital mortality in their regression analysis [26]. Peiffer et al., in their exploration of factors associated with adverse outcomes (e.g. prolonged length of stay, readmission, and mortality) following surgery in a Ghanaian regional hospital, found that although only 10% of the cohort were girls, female gender was significantly associated with in-hospital postoperative mortality [15*]. When looking at pediatric burn care in their investigation of potential relationships among patient demographics, thermal injury and mortality in a Malawi burn unit, Purcell et al. demonstrated that fewer females presented (44.1%), and that those who did were in more severe states (% total body surface area) [9*]. Perhaps due in part to the severity, the authors reported higher rates of surgical intervention for these girls, and interestingly, a lack of increased odds of mortality due to female gender [9*]. Allorto et al., while investigating factors associated with delayed children’s surgery and mortality following burn injuries in a South African burn service, reported fewer females in their cohort (44.5%); yet they were less likely to undergo early surgical intervention, they had higher mortality, and lengths of stay [12*]. Pediatric ophthalmology studies have identified a significant male predominance at presentation. For example, in reports concerning the proportions of children presenting for cataract surgery in a Nigerian ophthalmology clinic, Ugalahi et al. noted the paltry number of girls (44.4%) [31]. In their evaluation of several African child eye health tertiary facilities (CEHTFs), Adhisesha Reddy et al. found access disparities favouring boys for children receiving cataract surgery and follow up; they also noted little effective means to reduce the inequity [32]. Courtright et al., in their analysis of child recipients of cataract surgery at two Tanzanian CEHTFs found a sex ratio of 1.48 [33]. Eriksen et al. reported few girls (36.4%) in their examination of an active surgical service for children in a Tanzanian tertiary eye care referral facility [34]. They also found girls were less likely to attend follow up visits [34].

When discussing pediatric surgical disease, it must be acknowledged that there are certain diseases that affect boys more than girls, and vice versa. For example, congenital inguinal hernia is a very common pediatric condition, for which its surgical repair has been described in the literature as having a sex ratio prevalence (male:female) ranging from 2 to 18:1 [35, 36]. In Africa specifically, Igwe et al. reported a sex ratio of 5:1 in their comparison of laparoscopic versus open inguinal hernia repair, in a Nigerian university teaching hospital [37]. In addition, trauma may occur more often in males than females, as noted by Aluisio et al. and Halawa et al. in their Rwandan and Egyptian reports [38, 39]. Nevertheless, this current study excluded conditions that affected predominantly boys or girls, as a result of unique anatomy or physiology. As indicated in the methods definitions and caveats sub-section, this study incorporated conditions with equal sex ratios, meaning they occurred equally in boys and girls.

In this systematic review and meta-analysis of sex-disaggregated studies it was demonstrated that surgical interventions were more likely to be performed on males, and that females were more likely to have poor postoperative outcomes, including death. These findings are based on an analysis of 54 papers, mostly retrospective and single-site in nature, focusing on pediatric cohorts of up to 200 children, stemming from four African countries, mostly focusing on pediatric surgery. The male predominance in surgical access, and female predominance in adverse postoperative outcomes including mortality, were highlighted in sex ratio calculations, then reinforced through forest plot odds ratios in the meta-analysis.

There are 2 types of models for a meta-analysis: a fixed and random effects model. The latter was more appropriate for this study, because it assumed that the true effect could vary by study due to the differences among them (heterogeneity) [40, 41]. For example, the included studies varied by the size of their cohorts, location, types of surgical intervention, and other demographics. This could have resulted in different effect sizes. In the meta-analysis, there was high heterogeneity when evaluating access, and moderate heterogeneity when examining postoperative mortality. As a result, a prediction interval was calculated, and the vast majority of studies’ true effects were positive. The results were also supported by both the lack of statistical significance for statistical indices, and the absence of funnel plot asymmetry, both refuting publication bias.

In contrast to the Cochrane Risk of Bias tool (RoB2) [42], the recommended tool for randomized control trials, the Risk of Bias in Non-Randomized Studies – of Interventions (ROBINS-I) tool is used to evaluate the quality of the evidence for non-randomized studies [25]. In this study, only 22 of the studies were eligible for ROBINS-I assessment. These studies’ risk of bias level by domain and intervention were described (risk of bias assessment results sub-section) and illustrated (Table 2).

Unfortunately all of the studies assessed had a risk of bias in at least one domain, which limited the strength of the result interpretation. Within the domain for confounding, there was both critical and serious risk of bias in approximately 50% of the studies, which contributed to serious risk of bias across the pre-/at-intervention domains. As a result, the possibility of at least one domain not being appropriately measured or controlled cannot be excluded - nor can the possibility of confounding be entirely overlooked.

This study is a systematic review, providing a snapshot of the literature located in selected databases, published from specific African countries, during a discrete period of time. Although the analysis of publication bias was not significant according to the funnel plot (and associated statistical indices), the included studies represented approximately a third of all the countries in Africa. This study was a summary of what was reported. At no point did this study place judgment on African values, their family unit, their healthcare providers’ intentions or capabilities. Moreover, this study should not be viewed as an indictment of Africa’s treatment of its children, as similar findings have been reported in high-income countries. For example, in their thirteen year review of the epidemiology of pediatric surgery in the US, Rabbitts et al. reported female gender among other factors associated with decreased likelihood of surgical intervention on multivariate analysis [43]. In their examination of social determinants on pediatric trauma outcomes in the US South, Dickens et al. reported that girls had lower odds of being referred for rehabilitation services, and Black girls lowest of all groups [44]. In their analysis of access to renal transplantation, through a multinational European collaboration, Hogan et al. reported girls had lower probability of receiving preemptive transplantation, for reasons only partially attributed to medical factors [45]. In essence, what cannot be overstated is that findings of gender differentials in surgical access and/or postoperative outcomes have similarly been reported in high income countries; so gender disparities are not solely specific to Africa.

Other study limitations include: generalizability, the small number of included studies, the quality of data they presented, their age, and potential confounders (e.g. geographical location of participants, lack of infrastructure, etc). Studies with potential risk of bias were not removed from the analysis, nor was the one study that may have been a highly influential outlier.

This study was conducted in constant consultation with several African collaborators, who have attained positions of considerable distinction in pediatric surgery. In terms of the next phase in this study, collaboration with these colleagues and other African stakeholders (e.g. healthcare providers, caregivers, former patients, etc), is underway to uncover the obstacles to gender equity in pediatric surgical care, and to develop a comprehensive, sustainable mitigation plan to reduce the disparities via a mixed methods approach in Africa.

## Conclusion

African girls are a vulnerable population who, in addition to facing the limitations of residing in many low-resource settings, may be subjected to additional barriers of gender inequity. This study’s findings suggest that although surgeries are performed more often on males, females have a higher likelihood of adverse postoperative outcomes. This first-time summary of evidence of gender disparities in pediatric surgical access and outcomes in Africa is an urgent call to action, hopefully prompting not only further investigation, but above all tangible change.

## Disclosures

## Author Contributions

All authors contributed to the study conception and design. Material preparation, data collection and analysis was performed by SW, DP. The first draft of the manuscript was written by SW, and all authors edited, read, and approved the final manuscript.

## Data Availability

All data produced in the present study are available upon reasonable request to the authors.

## Appendix A Search Strategy

### Databases Searched

#### Africa-Wide Information [EBSCO] (March 31, 2022)

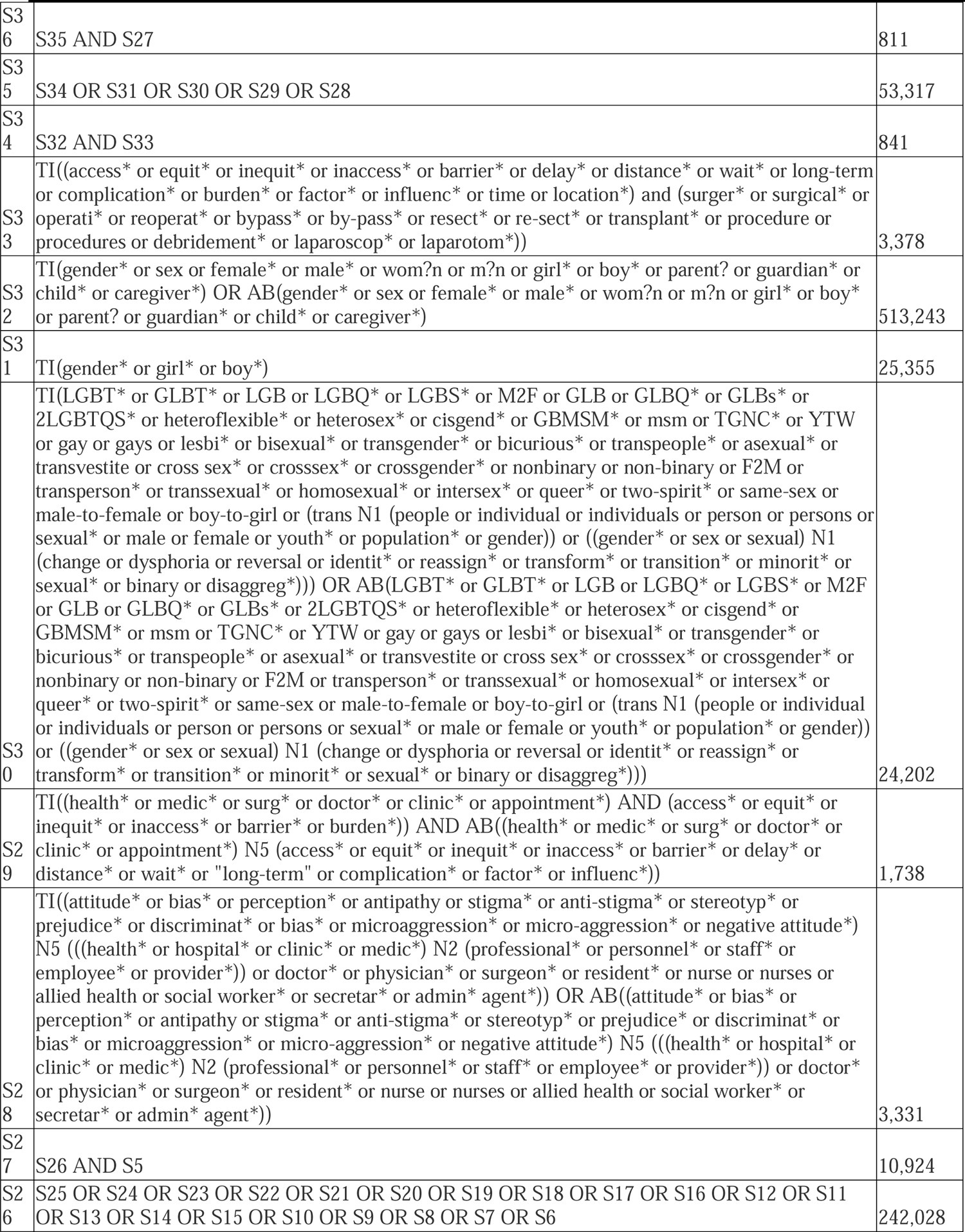
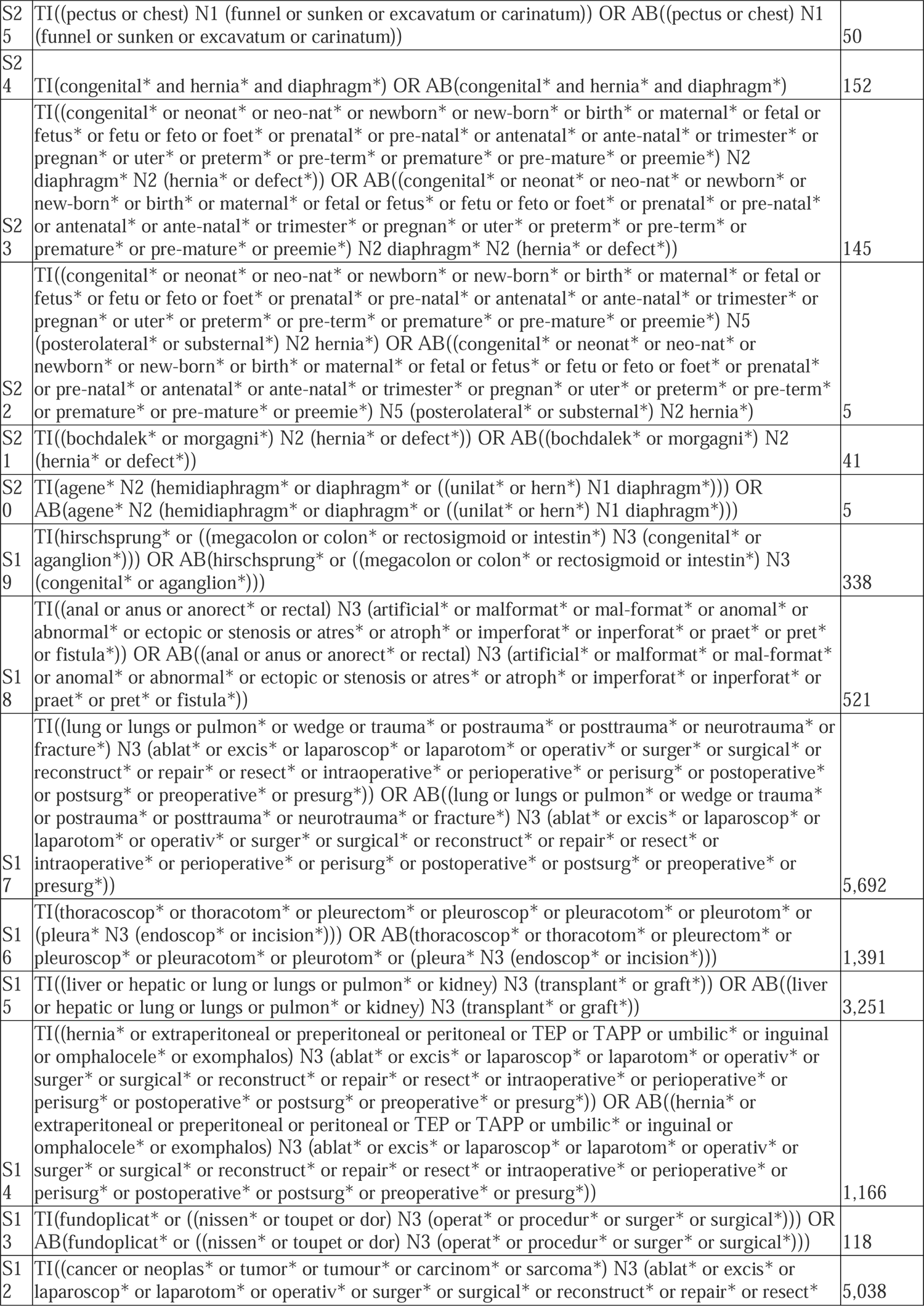
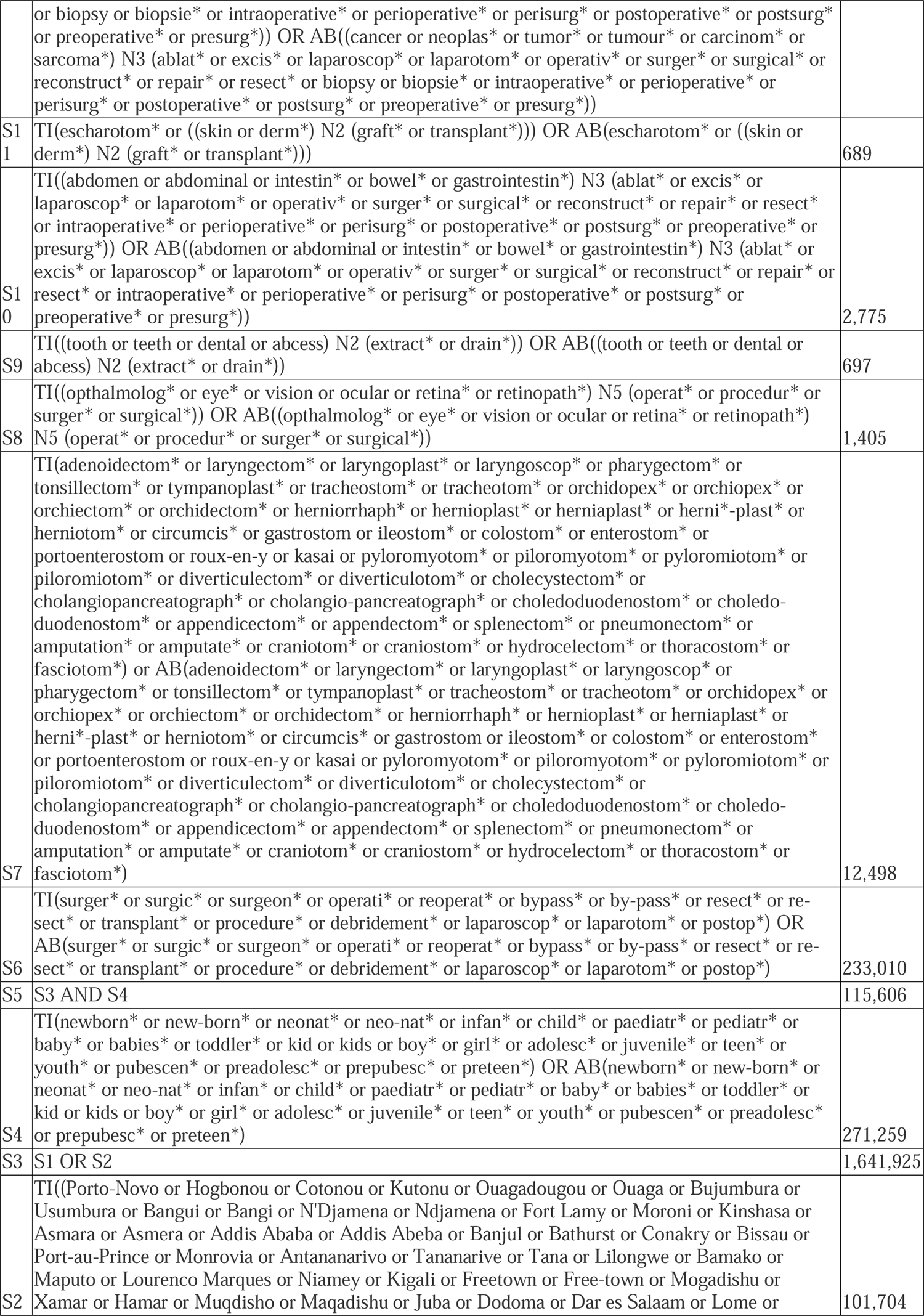
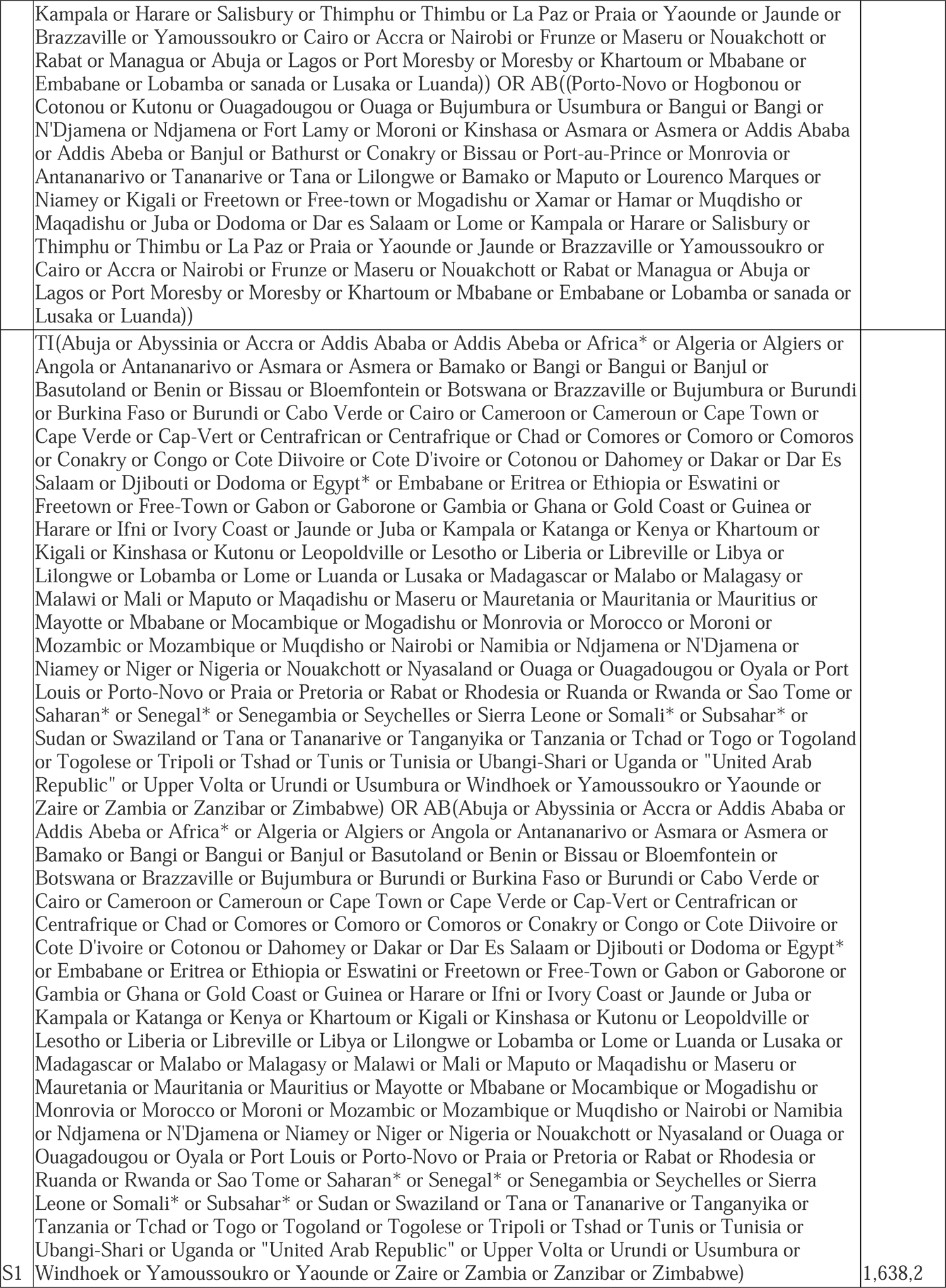

#### CINAHL Plus [EBSCO] (March 31, 2022)

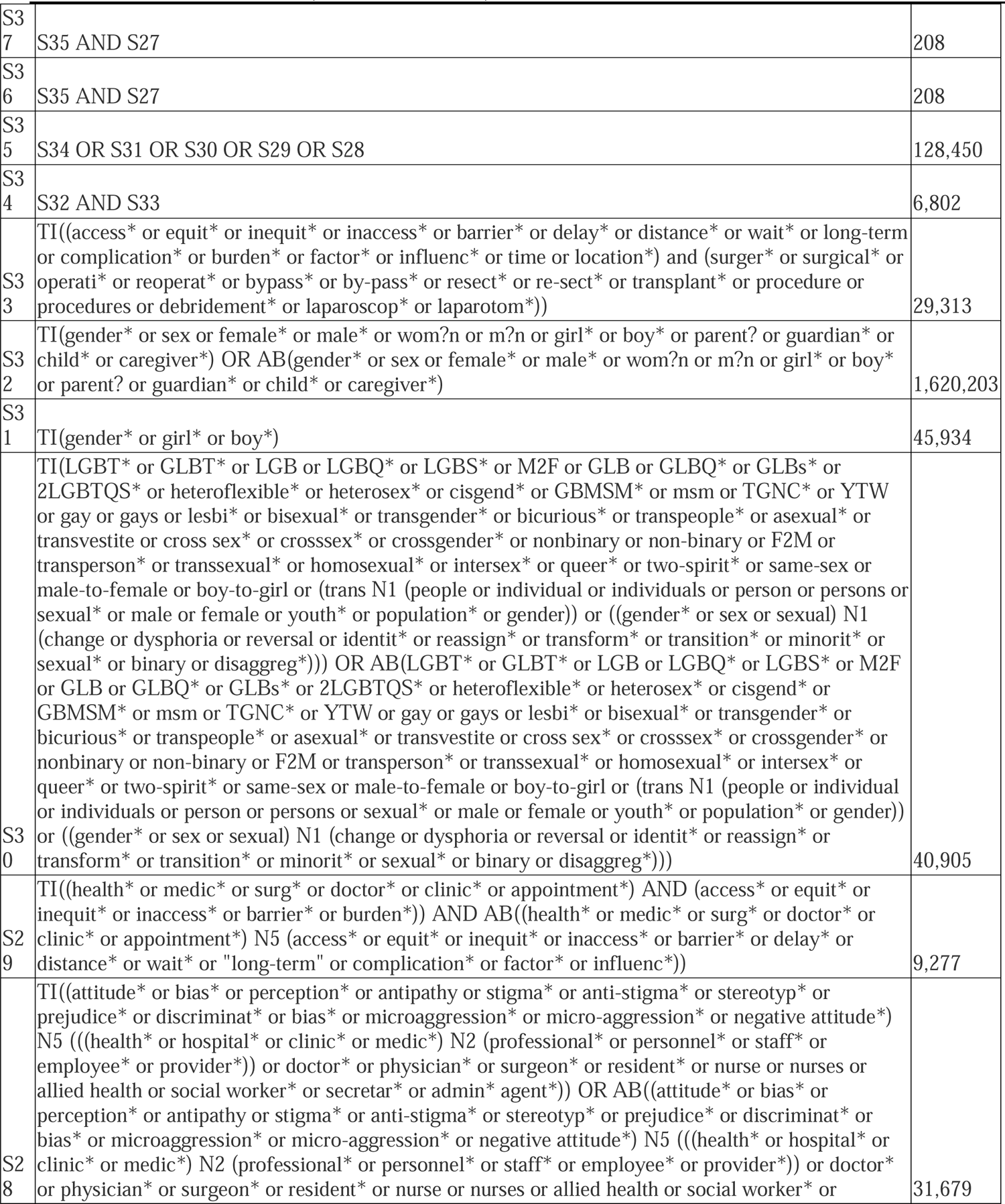
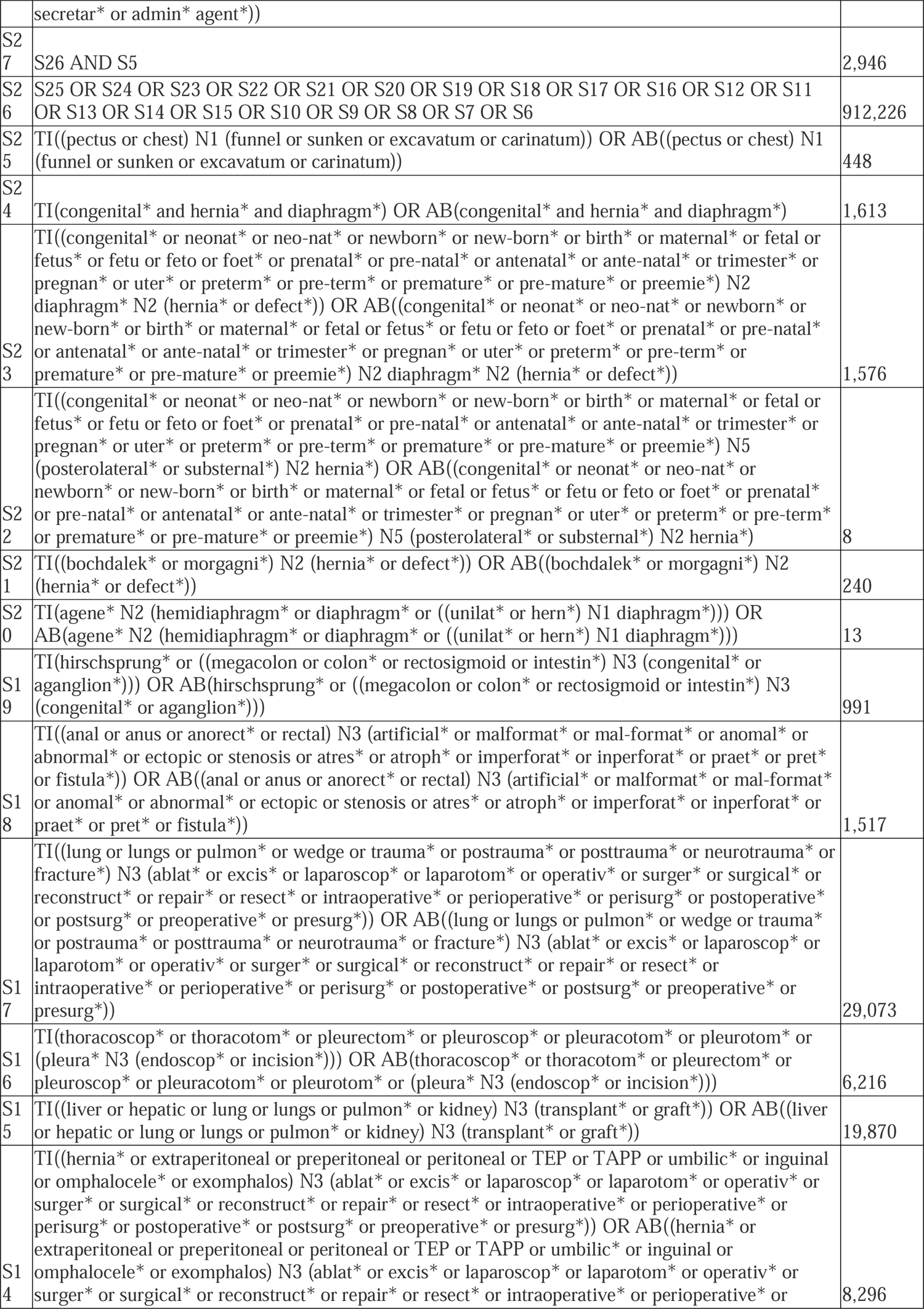
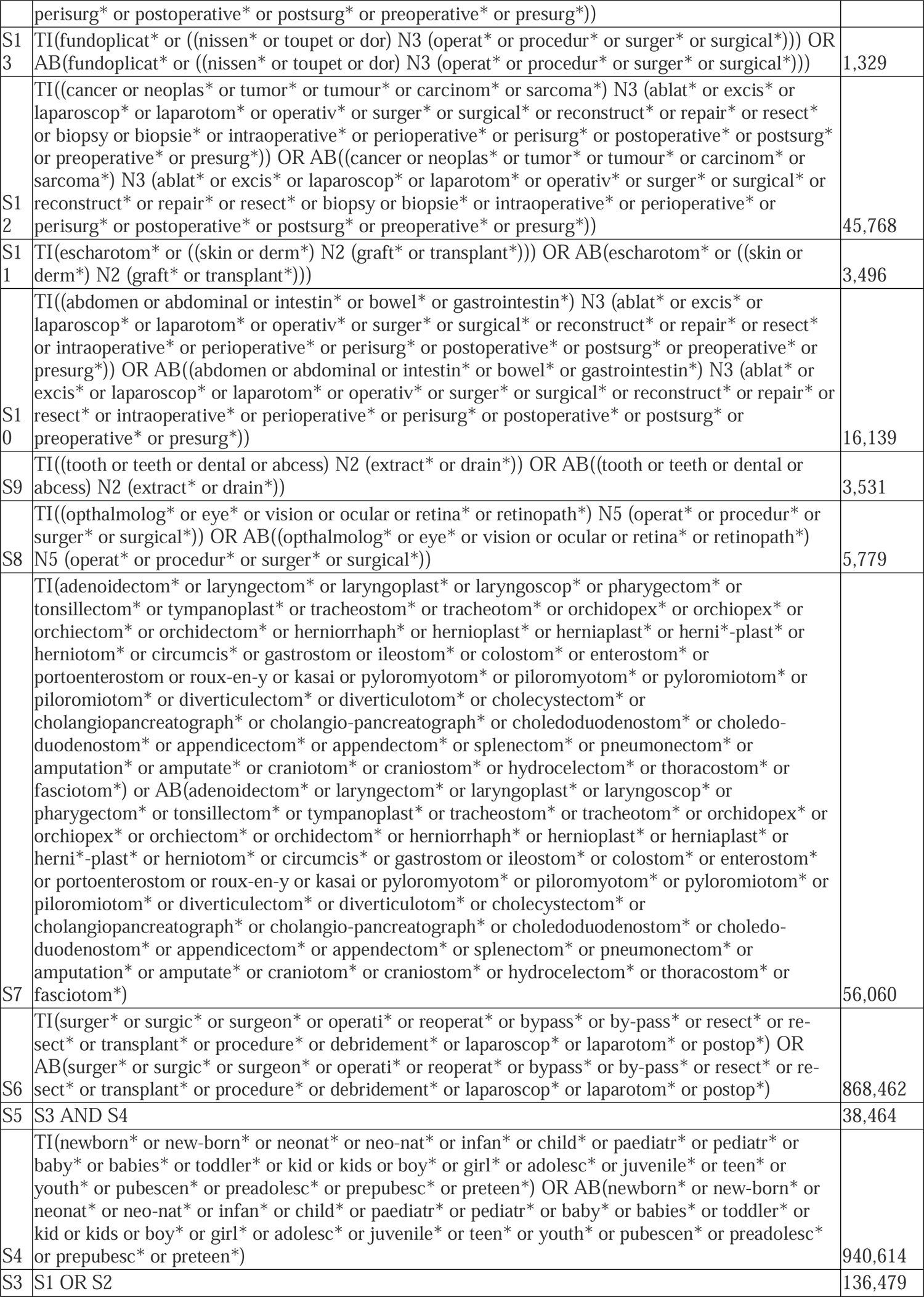
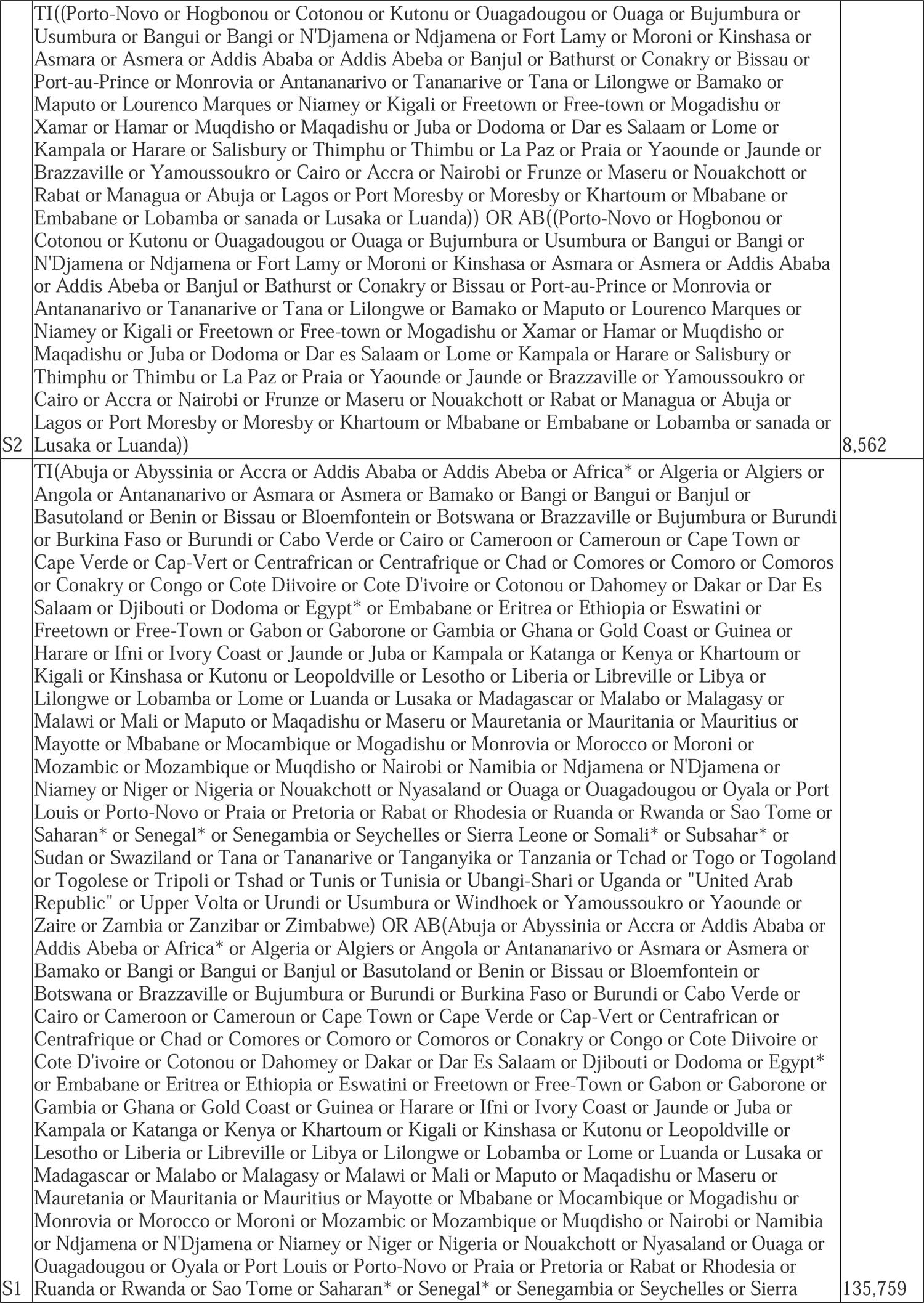
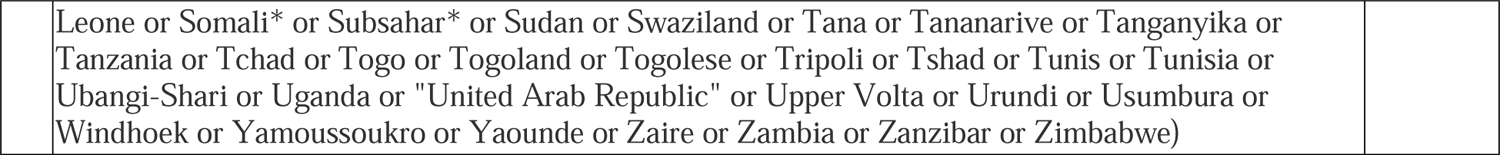

#### Cochrane [Wiley] (March 31, 2022)

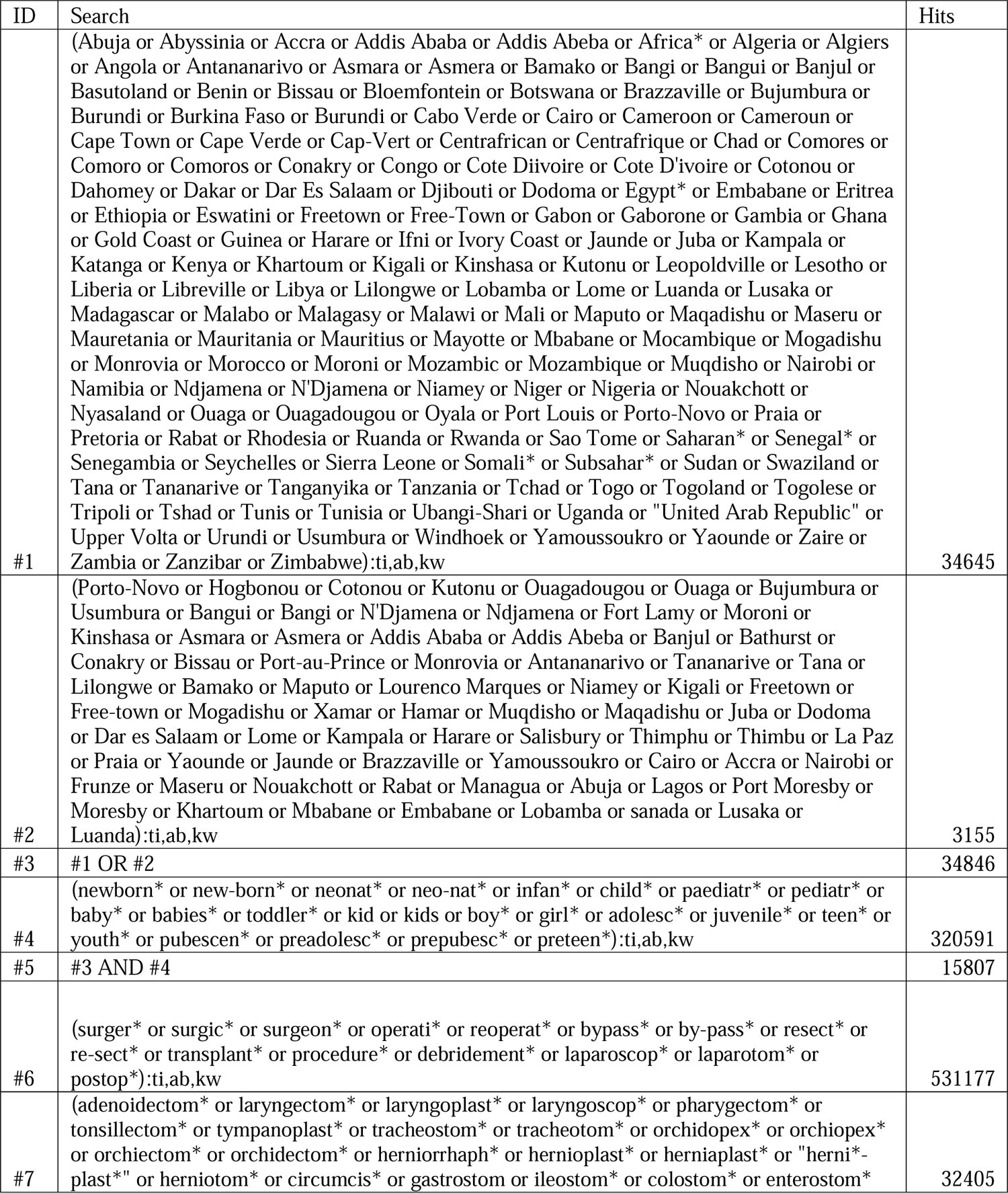
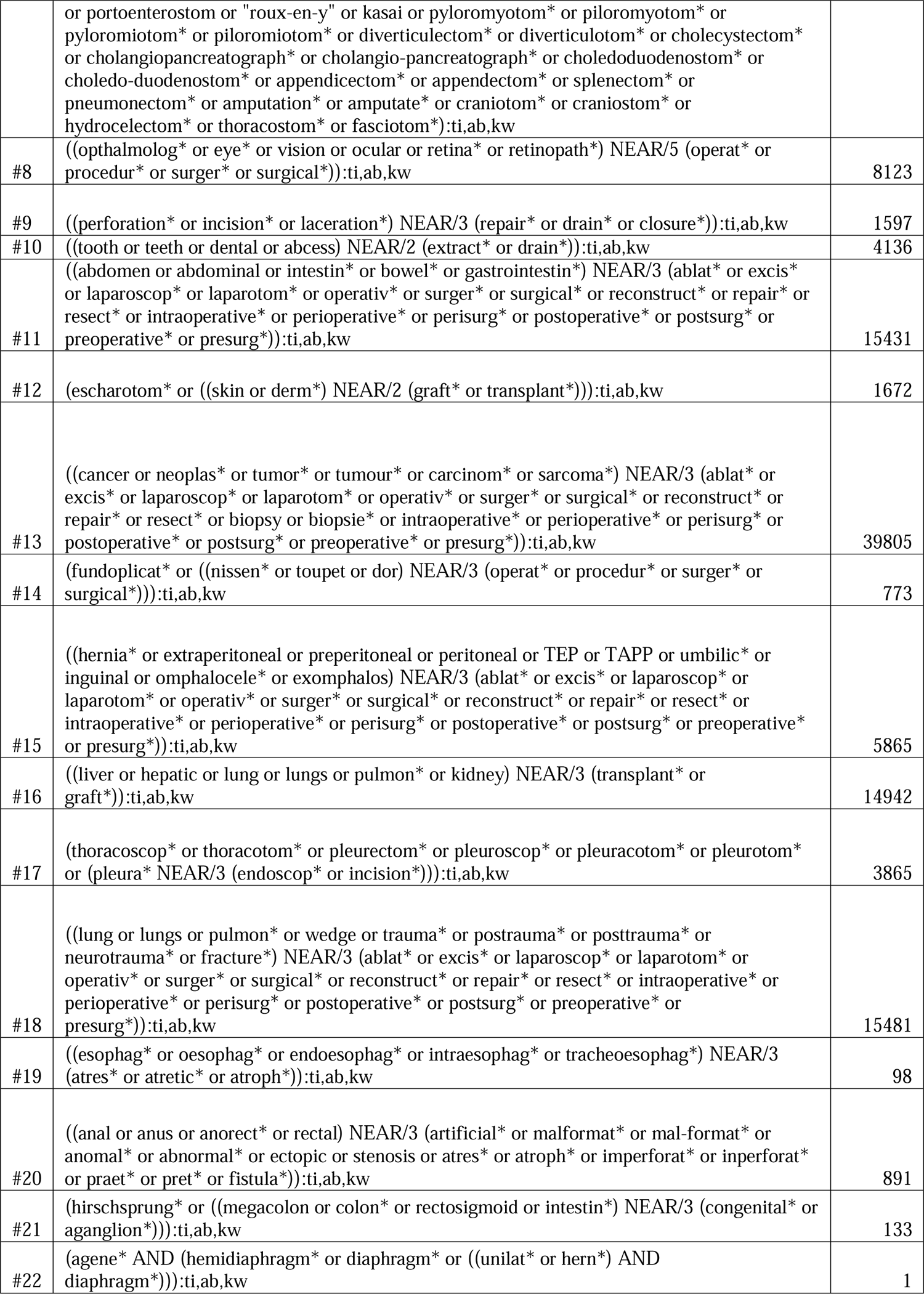
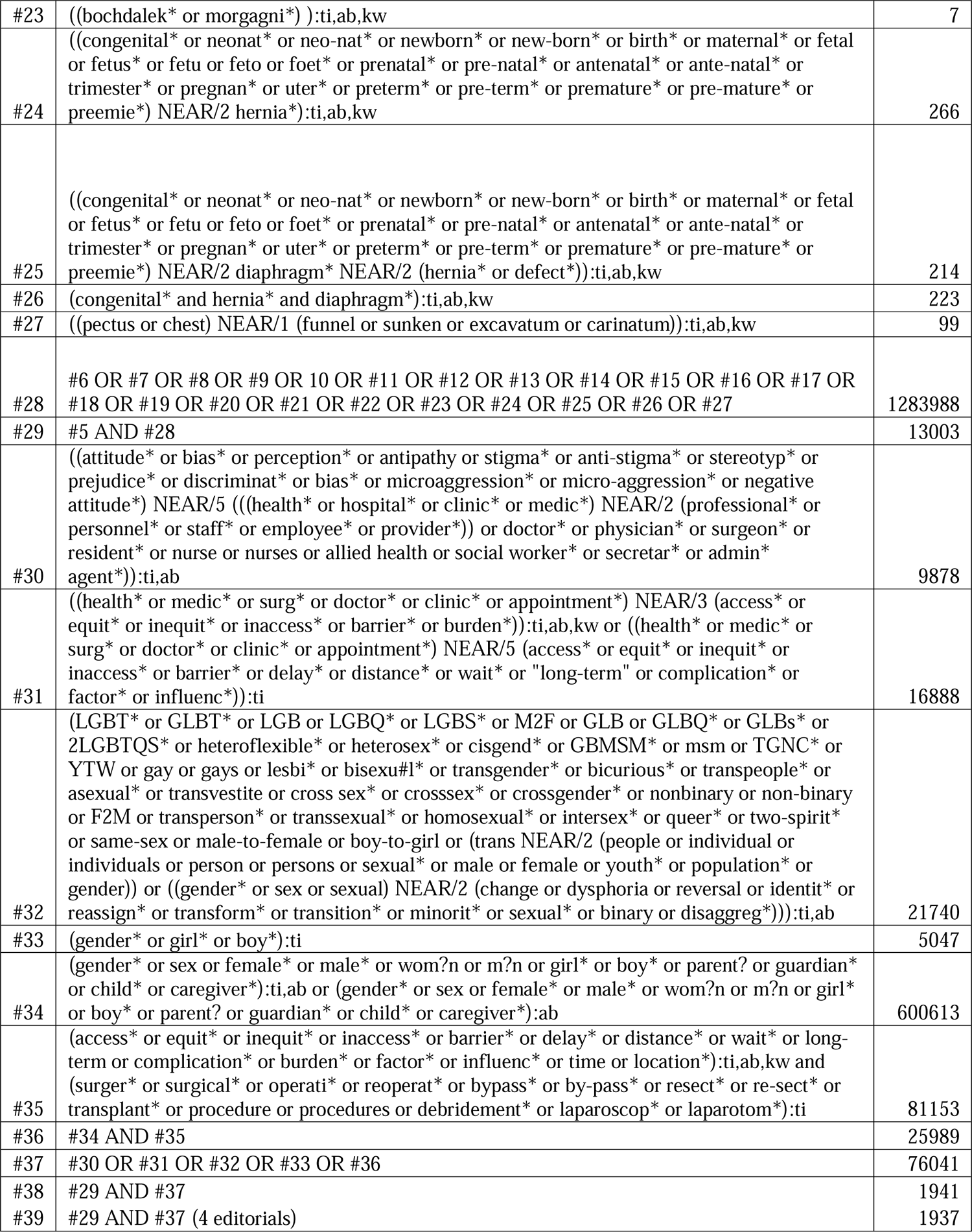

#### Embase [Ovid] (March 31, 2022)

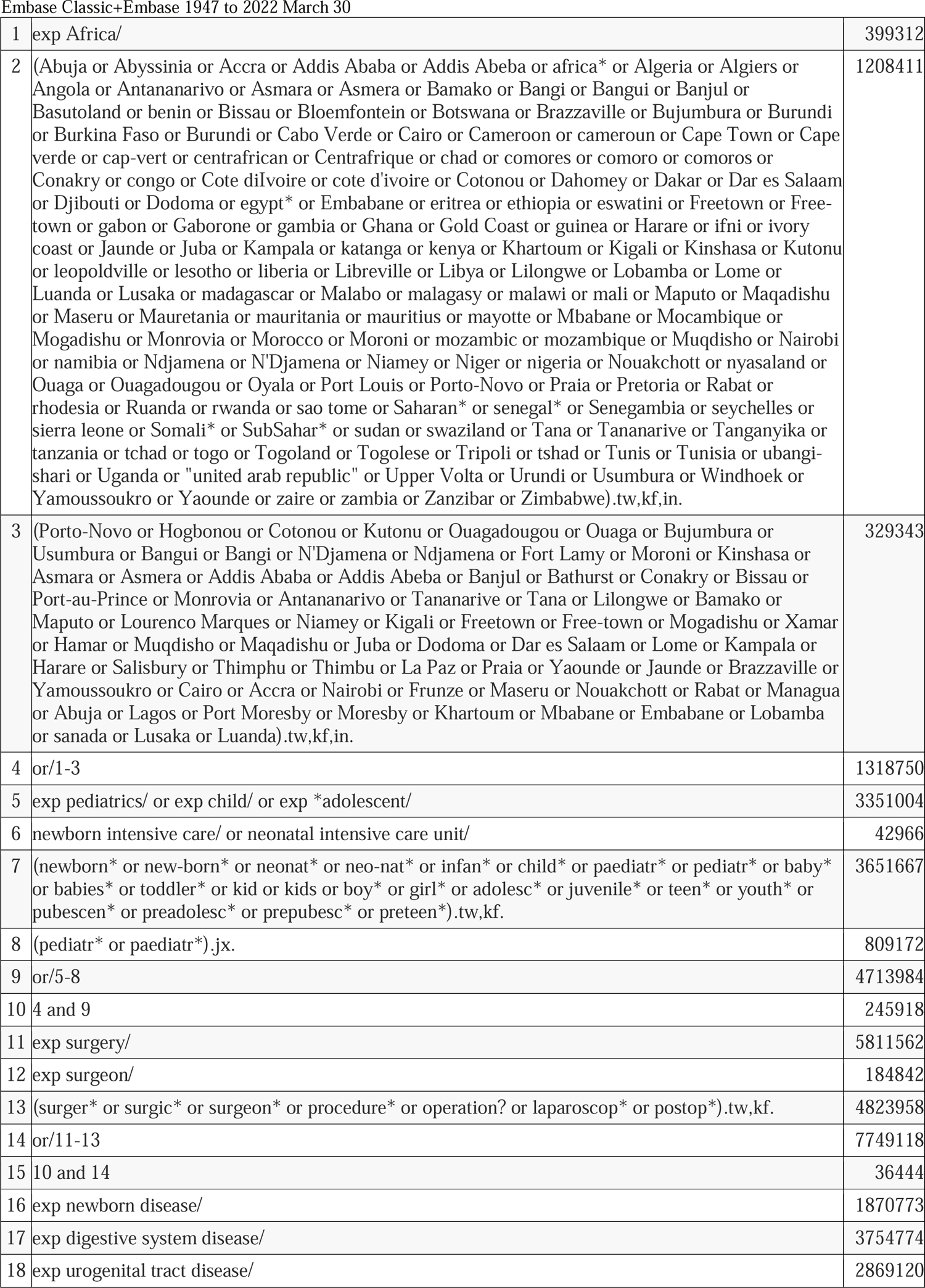
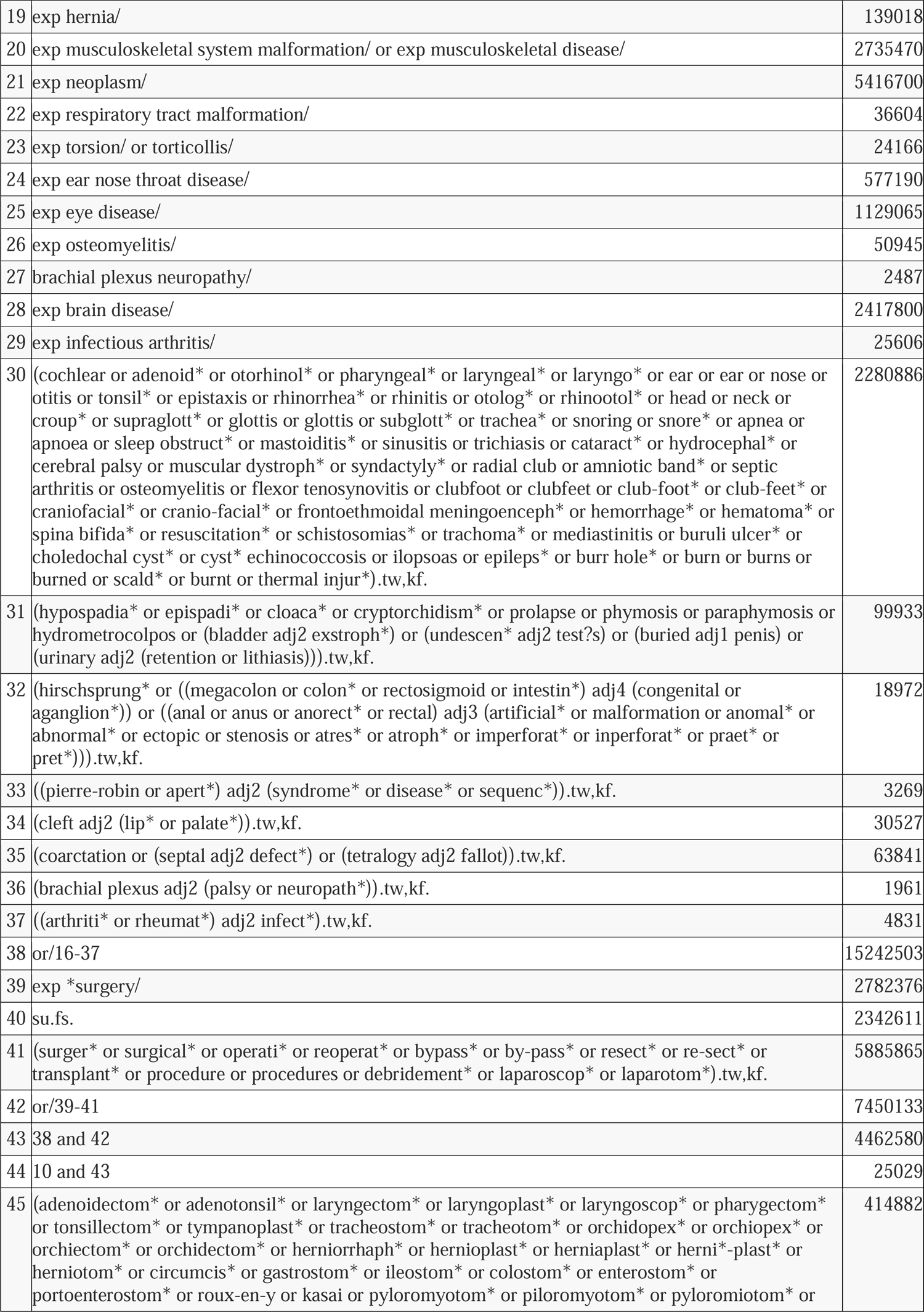
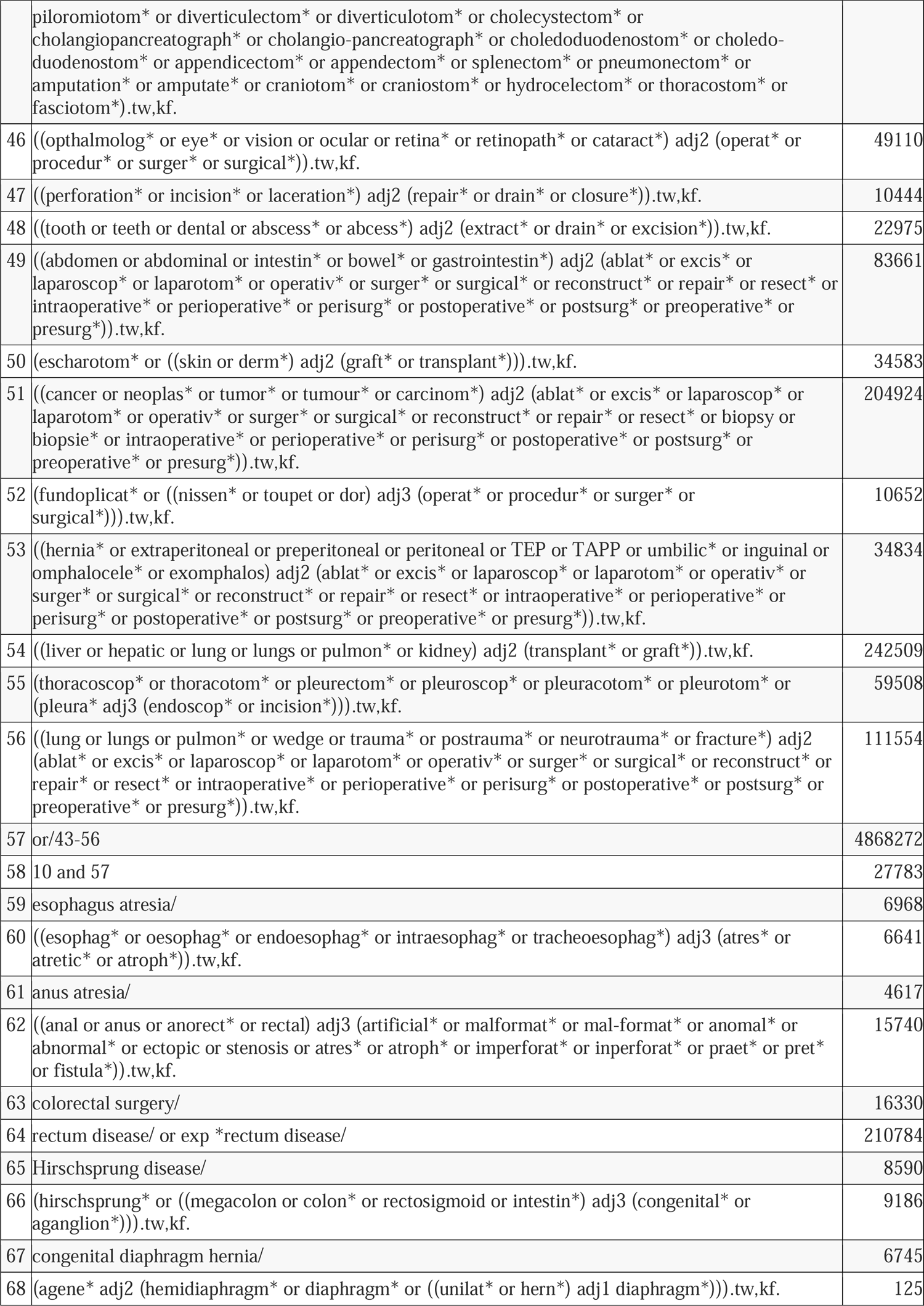
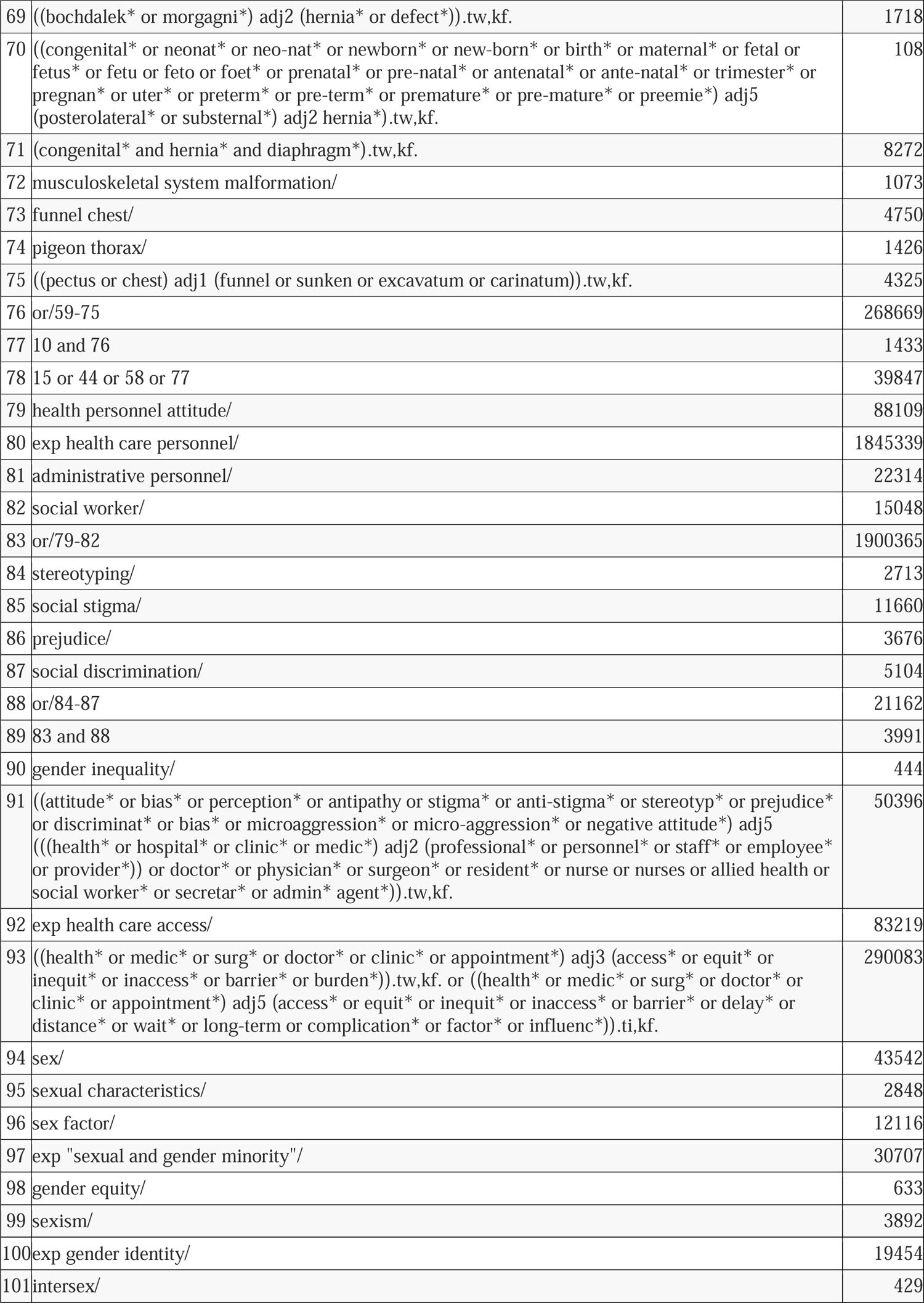
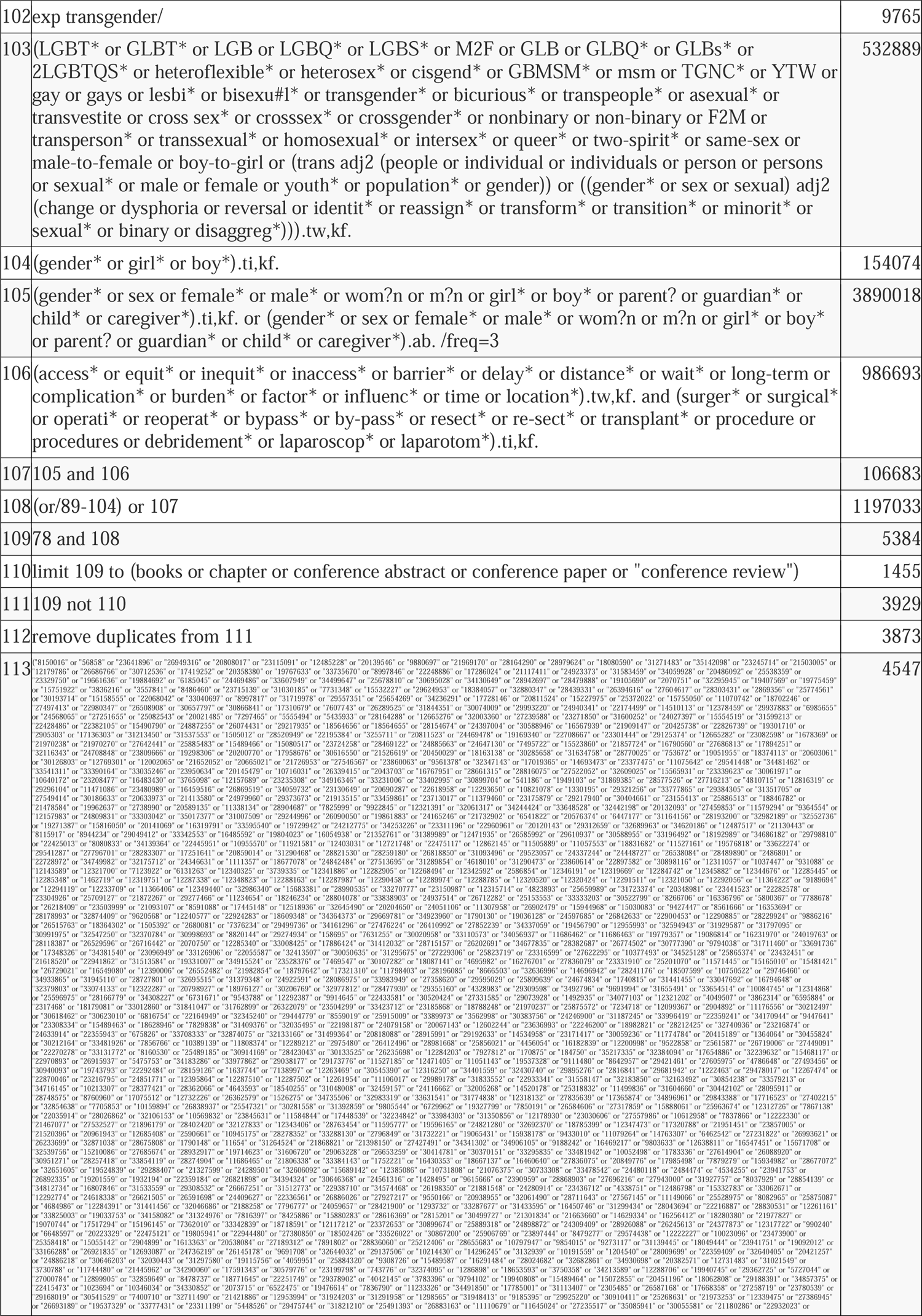
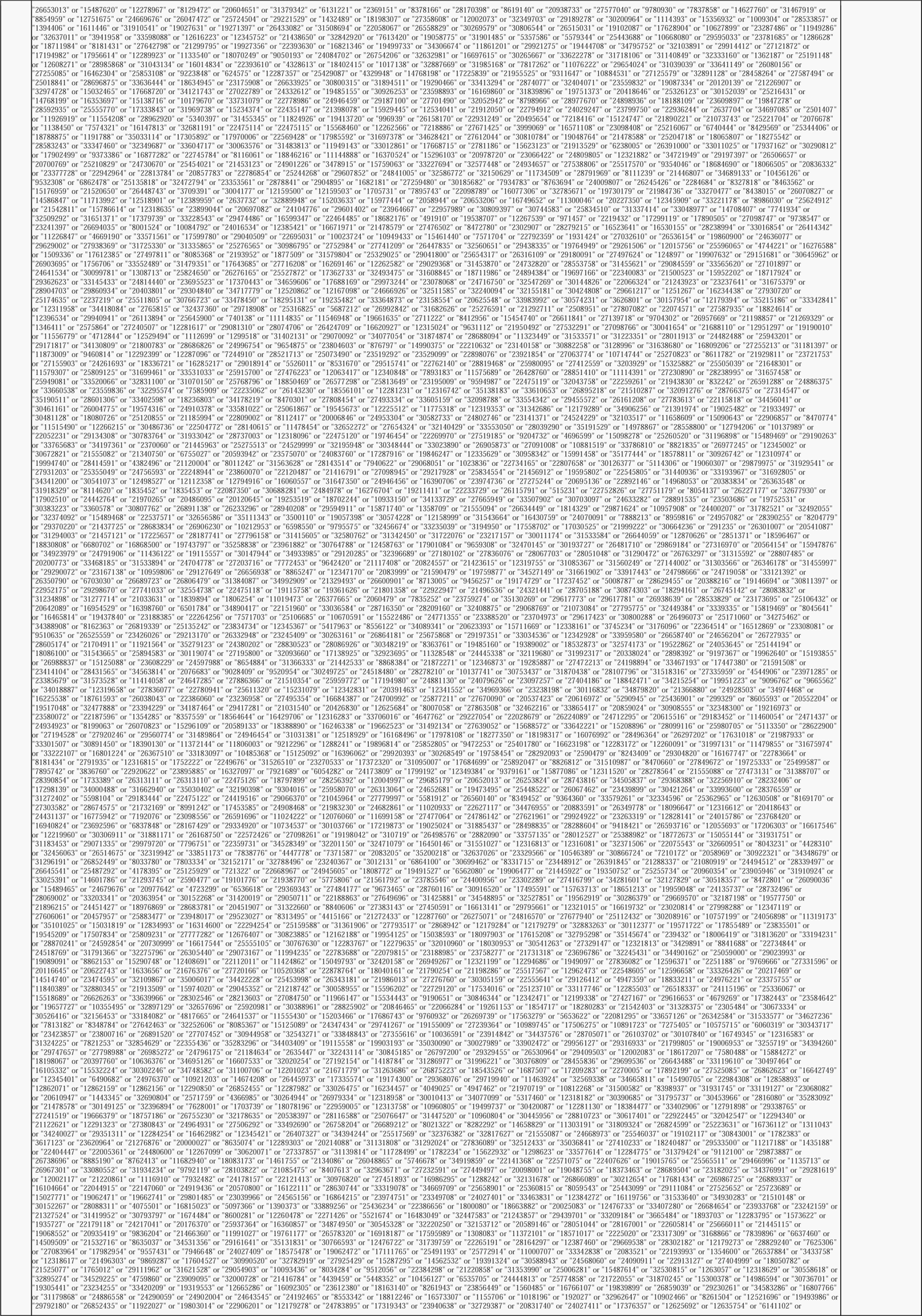
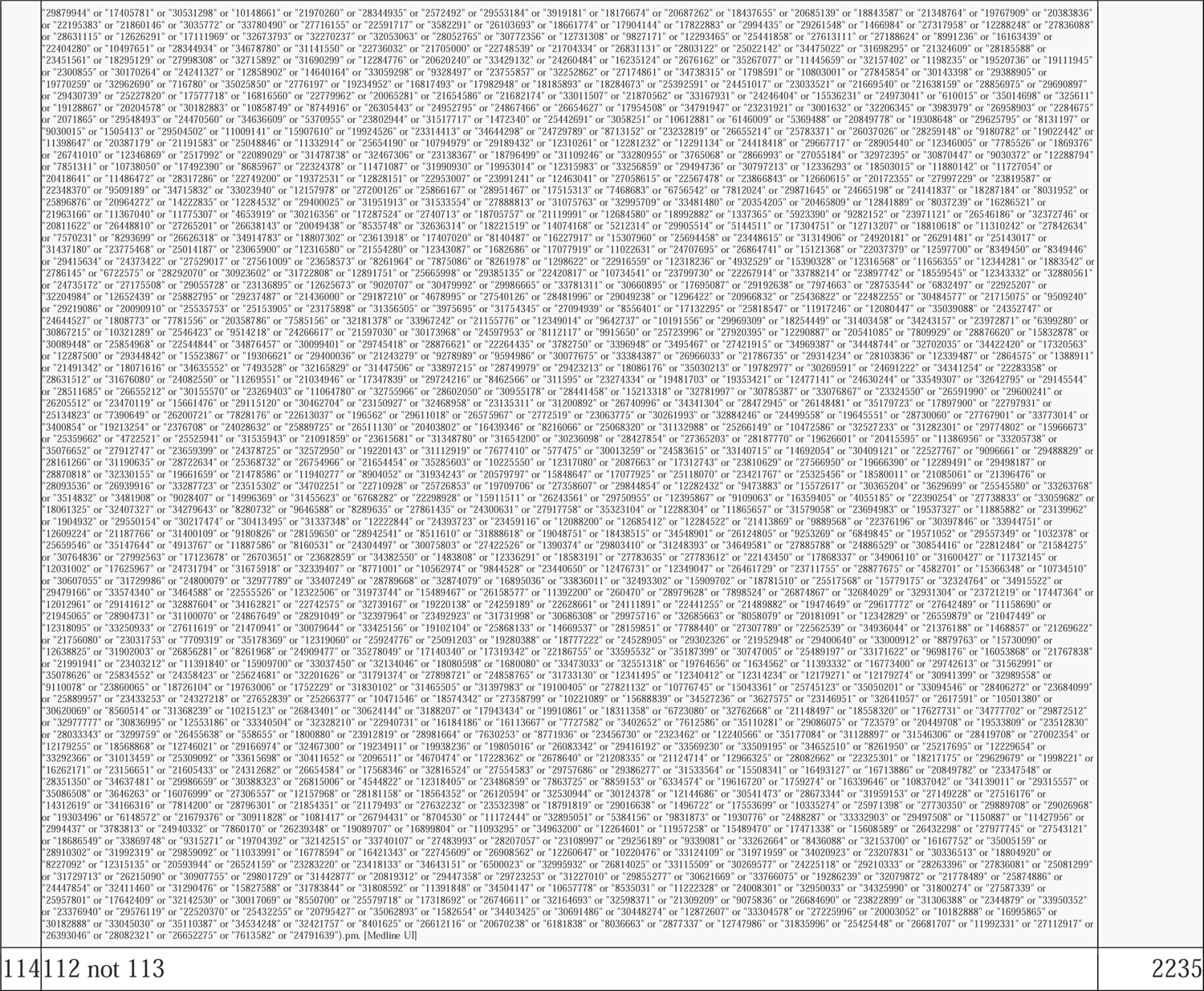

#### Global Health [Ovid] (March 31, 2022)

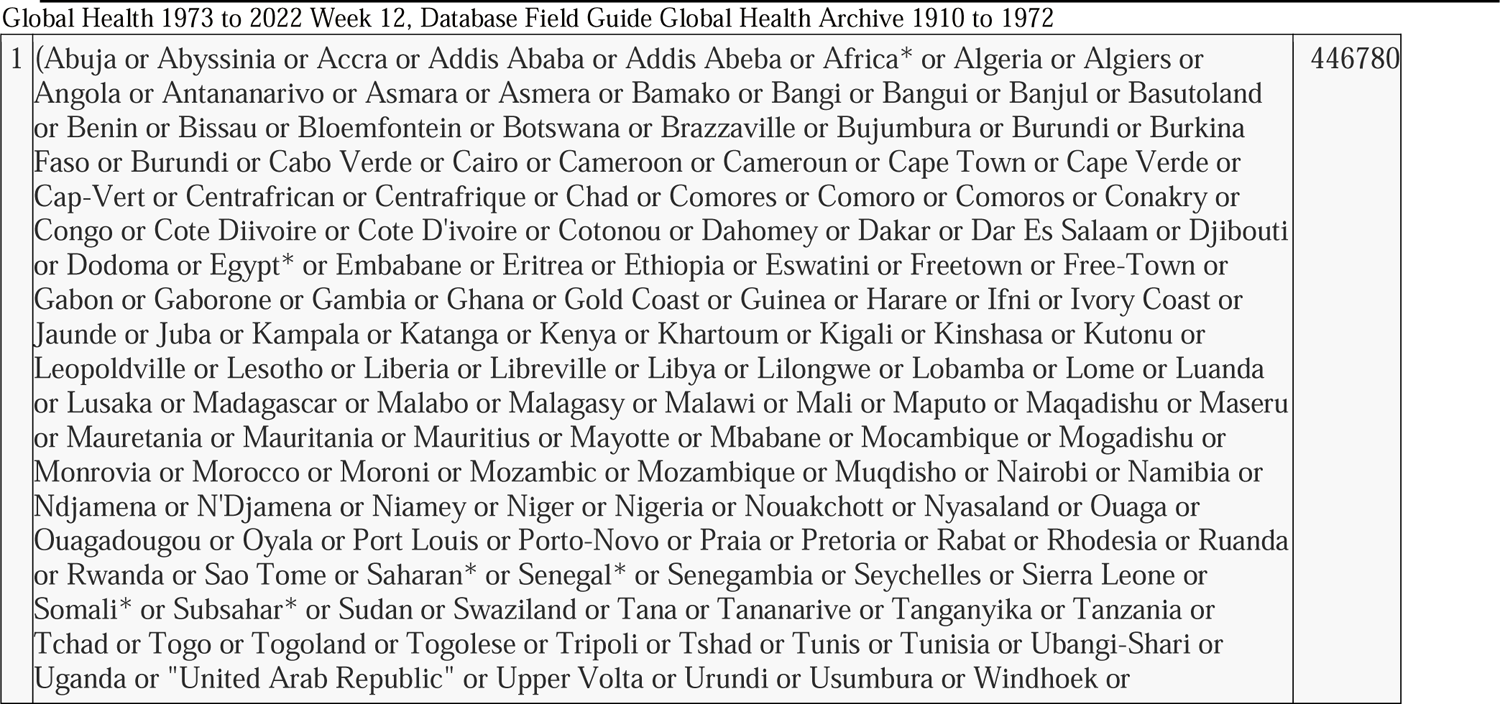
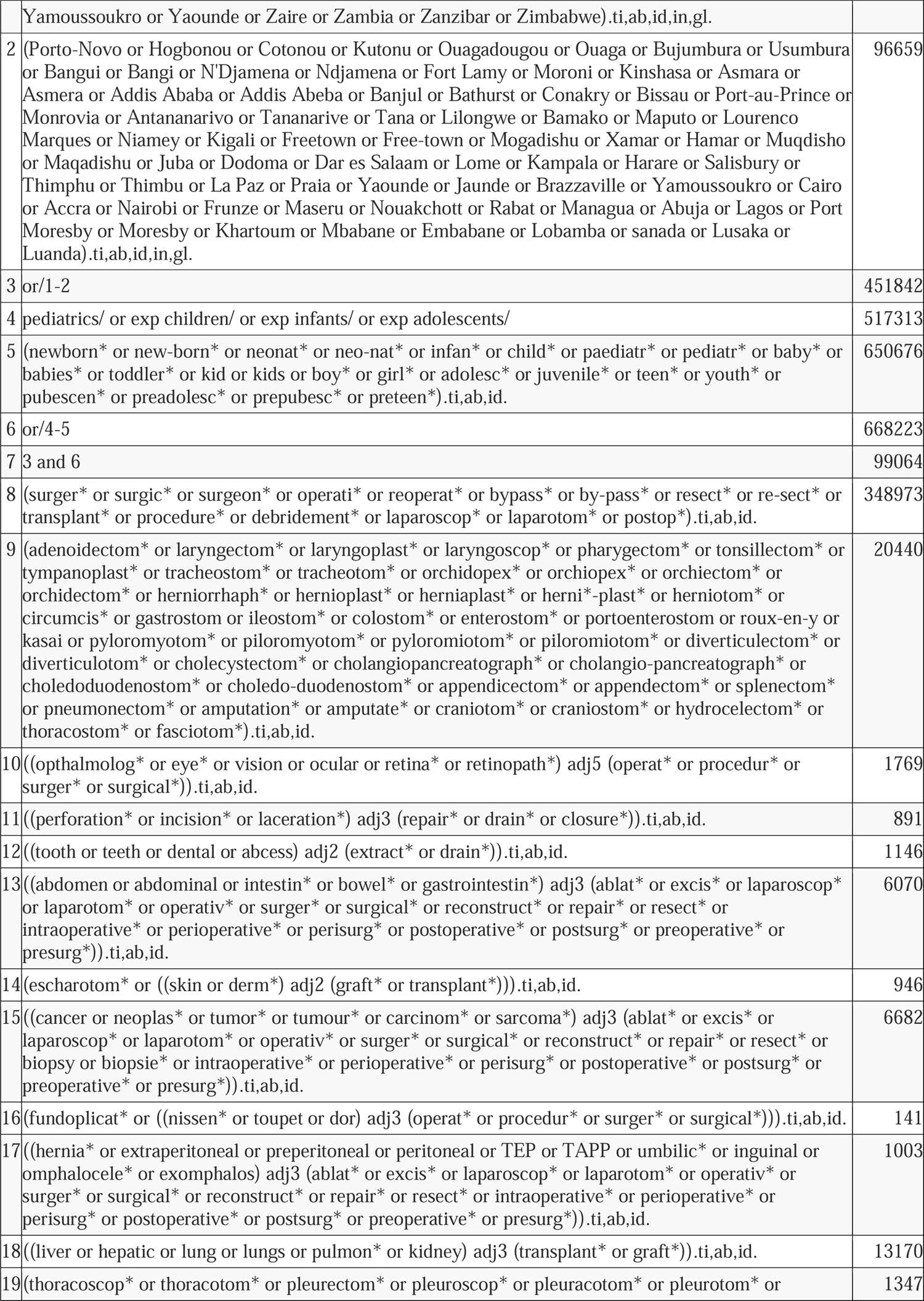
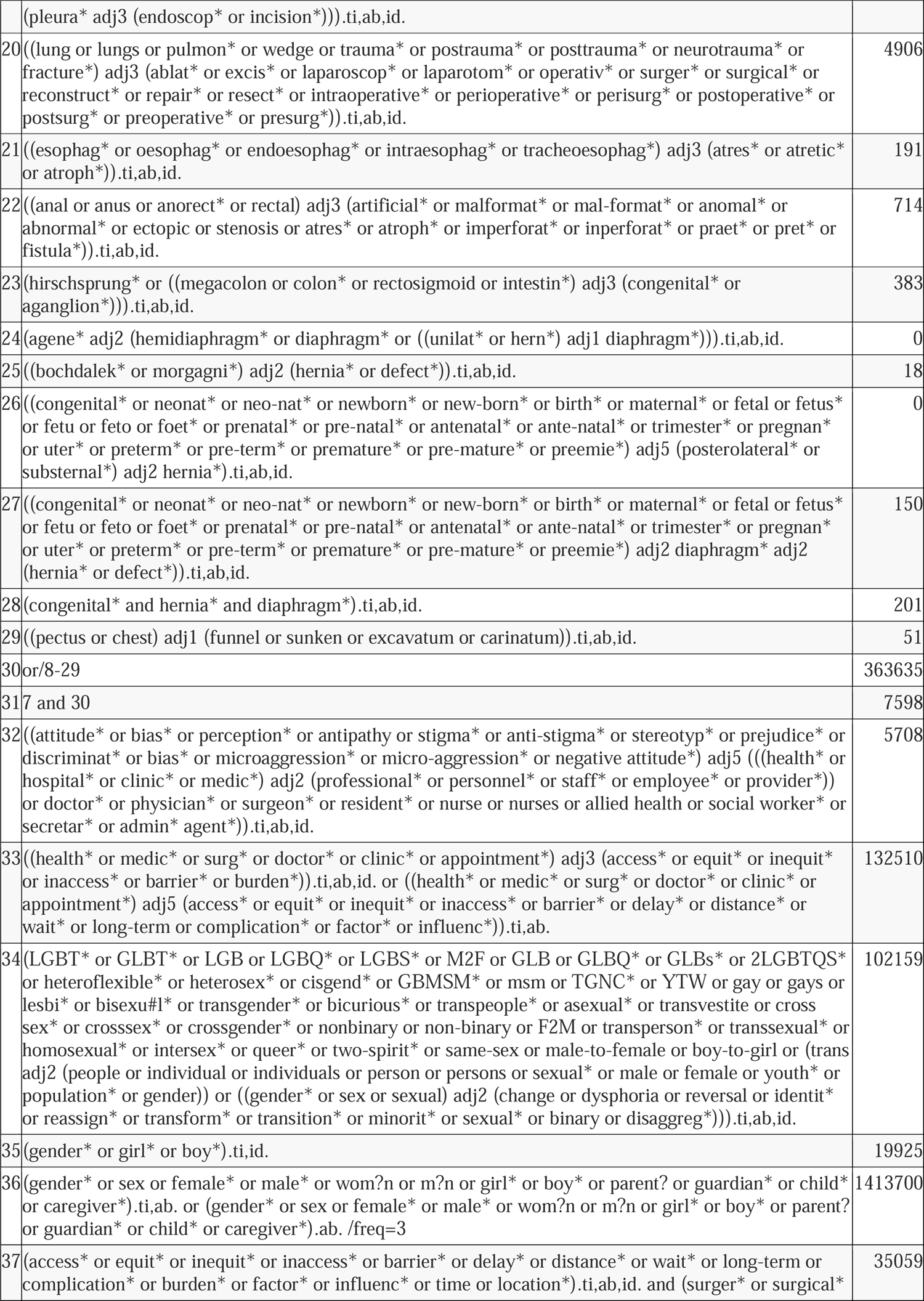
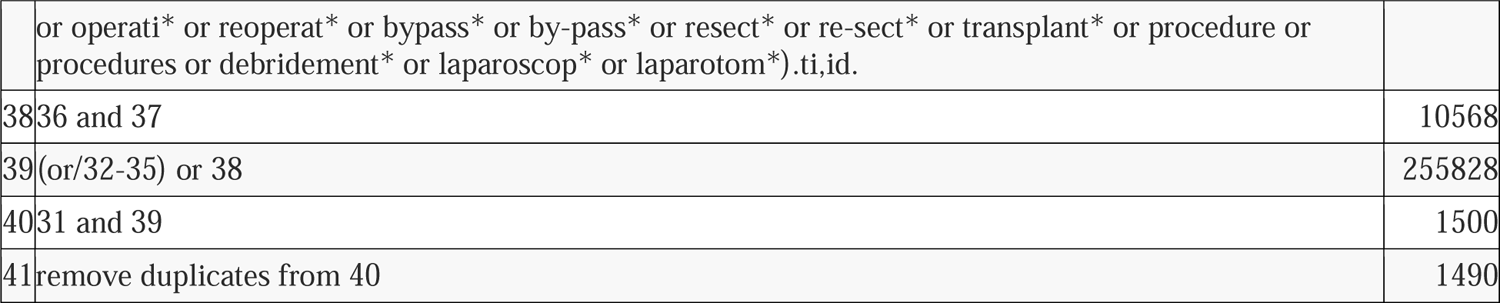

#### Global Index Medicus [WHO] (March 31, 2022)

|  |  |  |
| --- | --- | --- |
| 1 | (tw:((Abuja or Abyssinia or Accra or Addis Ababa or Addis Abeba or Africa* or Algeria or Algiers or Angola or Antananarivo or Asmara or Asmera or Bamako or Bangi or Bangui or Banjul or Basutoland or Benin or Bissau or Bloemfontein or Botswana or Brazzaville or Bujumbura or Burundi or Burkina Faso or Burundi or Cabo Verde or Cairo or Cameroon or Cameroun or Cape Town or Cape Verde or Cap-Vert or Centrafrican or Centrafrique or Chad or Comores or Comoro or Comoros or Conakry or Congo or Cote Diivoire or Cote D'ivoire or Cotonou or Dahomey or Dakar or Dar Es Salaam or Djibouti or Dodoma or Egypt* or Embabane or Eritrea or Ethiopia or Eswatini or Freetown or Free-Town or Gabon or Gaborone or Gambia or Ghana or Gold Coast or Guinea or Harare or Ifni or Ivory Coast or Jaunde or Juba or Kampala or Katanga or Kenya or Khartoum or Kigali or Kinshasa or Kutonu or Leopoldville or Lesotho or Liberia or Libreville or Libya or Lilongwe or Lobamba or Lome or Luanda or Lusaka or Madagascar or Malabo or Malagasy or Malawi or Mali or Maputo or Maqadishu or Maseru or Mauretania or Mauritania or Mauritius or Mayotte or Mbabane or Mocambique or Mogadishu or Monrovia or Morocco or Moroni or Mozambic or Mozambique or Muqdisho or Nairobi or Namibia or Ndjamena or N'Djamena or Niamey or Niger or Nigeria or Nouakchott or Nyasaland or Ouaga or Ouagadougou or Oyala or Port Louis or Porto-Novo or Praia or Pretoria or Rabat or Rhodesia or Ruanda or Rwanda or Sao Tome or Saharan* or Senegal* or Senegambia or Seychelles or Sierra Leone or Somali* or Subsahar* or Sudan or Swaziland or Tana or Tananarive or Tanganyika or Tanzania or Tchad or Togo or Togoland or Togolese or Tripoli or Tshad or Tunis or Tunisia or Ubangi-Shari or Uganda or "United Arab Republic" or Upper Volta or Urundi or Usumbura or Windhoek or Yamoussoukro or Yaounde or Zaire or Zambia or Zanzibar or Zimbabwe or Porto-Novo or Hogbonou or Cotonou or Kutonu or Ouagadougou or Ouaga or Bujumbura or Usumbura or Bangui or Bangi or N'Djamena or Ndjamena or Fort Lamy or Moroni or Kinshasa or Asmara or Asmera or Addis Ababa or Addis Abeba or Banjul or Bathurst or Conakry or Bissau or Port-au-Prince or Monrovia or Antananarivo or Tananarive or Tana or Lilongwe or Bamako or Maputo or Lourenco Marques or Niamey or Kigali or Freetown or Free-town or Mogadishu or Xamar or Hamar or Muqdisho or Maqadishu or Juba or Dodoma or Dar es Salaam or Lome or Kampala or Harare or Salisbury or Thimphu or Thimbu or La Paz or Praia or Yaounde or Jaunde or Brazzaville or Yamoussoukro or Cairo or Accra or Nairobi or Frunze or Maseru or Nouakchott or Rabat or Managua or Abuja or Lagos or Port Moresby or Moresby or Khartoum or Mbabane or Embabane or Lobamba or sanada or Lusaka or Luanda))) AND (tw:(surger* OR surgic* OR operation? OR postop*)) AND (tw:(gender* or sex or female* or male* or wom?n or m?n or girl* or boy* )) AND (tw:(attitude* or bias* or perception* or antipathy or stigma* or anti-stigma* or stereotyp* or prejudice* or discriminat* or bias* or microaggression* or micro-aggression* or access* or equit* or inequit* or inaccess* or barrier* or delay* )) AND ( limit:("adolescent" OR "child" OR "child, preschool" OR "infant" OR "newborn")) | 316 |

#### Medline [Ovid] (March 31, 2022)

|  |  |  |
| --- | --- | --- |
| 1 | exp Africa/ | 300924 |
| 2 | (Abuja or Abyssinia or Accra or Addis Ababa or Addis Abeba or Africa* or Algeria or Algiers or Angola or Antananarivo or Asmara or Asmera or Bamako or Bangi or Bangui or Banjul or Basutoland or Benin or Bissau or Bloemfontein or Botswana or Brazzaville or Bujumbura or Burundi or Burkina Faso or Burundi or Cabo Verde or Cairo or Cameroon or Cameroun or Cape Town or Cape Verde or Cap-Vert or Centrafrican or Centrafrique or Chad or Comores or Comoro or Comoros or Conakry or Congo or Cote Diivoire or Cote D'ivoire or Cotonou or Dahomey or Dakar or Dar Es Salaam or Djibouti or Dodoma or Egypt* or Embabane or Eritrea or Ethiopia or Eswatini or Freetown or Free-Town or Gabon or Gaborone or Gambia or Ghana or Gold Coast or Guinea or Harare or Ifni or Ivory Coast or Jaunde or Juba or Kampala or Katanga or Kenya or Khartoum or Kigali or Kinshasa or | 588882 |
|  | Kutonu or Leopoldville or Lesotho or Liberia or Libreville or Libya or Lilongwe or Lobamba or Lome or Luanda or Lusaka or Madagascar or Malabo or Malagasy or Malawi or Mali or Maputo or Maqadishu or Maseru or Mauretania or Mauritania or Mauritius or Mayotte or Mbabane or Mocambique or Mogadishu or Monrovia or Morocco or Moroni or Mozambic or Mozambique or Muqdisho or Nairobi or Namibia or Ndjamena or N'Djamena or Niamey or Niger or Nigeria or Nouakchott or Nyasaland or Ouaga or Ouagadougou or Oyala or Port Louis or Porto-Novo or Praia or Pretoria or Rabat or Rhodesia or Ruanda or Rwanda or Sao Tome or Saharan* or Senegal* or Senegambia or Seychelles or Sierra Leone or Somali* or Subsahar* or Sudan or Swaziland or Tana or Tananarive or Tanganyika or Tanzania or Tchad or Togo or Togoland or Togolese or Tripoli or Tshad or Tunis or Tunisia or Ubangi-Shari or Uganda or "United Arab Republic" or Upper Volta or Urundi or Usumbura or Windhoek or Yamoussoukro or Yaounde or Zaire or Zambia or Zanzibar or Zimbabwe).tw,kf,ia. |  |
| 3 | (Porto-Novo or Hogbonou or Cotonou or Kutonu or Ouagadougou or Ouaga or Bujumbura or Usumbura or Bangui or Bangi or N'Djamena or Ndjamena or Fort Lamy or Moroni or Kinshasa or Asmara or Asmera or Addis Ababa or Addis Abeba or Banjul or Bathurst or Conakry or Bissau or Port-au-Prince or Monrovia or Antananarivo or Tananarive or Tana or Lilongwe or Bamako or Maputo or Lourenco Marques or Niamey or Kigali or Freetown or Free-town or Mogadishu or Xamar or Hamar or Muqdisho or Maqadishu or Juba or Dodoma or Dar es Salaam or Lome or Kampala or Harare or Salisbury or Thimphu or Thimbu or La Paz or Praia or Yaounde or Jaunde or Brazzaville or Yamoussoukro or Cairo or Accra or Nairobi or Frunze or Maseru or Nouakchott or Rabat or Managua or Abuja or Lagos or Port Moresby or Moresby or Khartoum or Mbabane or Embabane or Lobamba or sanada or Lusaka or Luanda).tw,kf,ia. | 36496 |
| 4 | or/1-3 [Africa] | 651001 |
| 5 | pediatrics/ or exp adolescent/ or exp child/ or exp infant/ or adolescent medicine/ | 3843208 |
| 6 | Intensive Care Units, Neonatal/ | 16975 |
| 7 | (newborn* or new-born* or neonat* or neo-nat* or infan* or child* or adolesc* or paediatr* or pediater* or baby* or babies* or toddler* or kid or kids or boy* or girl* or juvenile* or teen* or youth* or pubescen* or preadolesc* or prepubesc* or preteen*).tw,kf. | 2720333 |
| 8 | (pediatr* or paediatr*).jw. | 611543 |
| 9 | or/5-8 | 4712562 |
| 10 | 4 and 9 [Africa Peds] | 181586 |
| 11 | exp Specialties, Surgical/ | 212232 |
| 12 | exp Surgical Procedures, Operative/ | 3401688 |
| 13 | exp Surgeons/ | 13730 |
| 14 | su.fs. | 2150629 |
| 15 | (surger* or surgic* or surgeon* or procedure* or operation? or laparoscop* or postop*).tw,kf. | 3441466 |
| 16 | or/11-15 [Gen Surg] | 5752042 |
| 17 | 10 and 16 [Africa Peds Gen Surg] | 21640 |
| 18 | exp "Congenital, Hereditary, and Neonatal Diseases and Abnormalities"/ | 1314461 |
| 19 | exp Digestive system diseases/ | 1868152 |
| 20 | exp Urologic Diseases/ or exp Male urogenital diseases/ or exp Female urogenital diseases/ or exp Prolapse/ | 1497917 |
| 21 | exp Hernia/ | 80680 |
| 22 | exp Musculoskeletal Abnormalities/ or exp Musculoskeletal Diseases/ | 1160670 |
| 23 | exp Neoplasms/ | 3651975 |
| 24 | exp Respiratory System Abnormalities/ | 11924 |
| 25 | exp Torsion Abnormality/ or Torticollis/ | 13585 |
| 26 | exp Otorhinolaryngologic Diseases/ | 394603 |
| 27 | exp Eye Diseases/ | 609143 |
| 28 | exp Osteomyelitis/ | 23926 |
| 29 | exp Brachial Plexus Neuropathies/ | 4077 |
| 30 | exp Hemorrhage/ | 350014 |
| 31 | exp Brain Diseases/ | 1331460 |
| 32 | exp Arthritis, Infectious/ | 15317 |
| 33 | (cochlear or adenoid* or otorhinol* or pharyngeal* or laryngeal* or laryngo* or ear or ear or nose or otitis or tonsil* or epistaxis or rhinorrhea* or rhinitis or otolog* or rhinootol* or head or neck or croup* or supraglott* or glottis or glottis or subglott* or trachea* or snoring or snore* or apnea or apnoea or sleep obstruct* or mastoiditis* or sinusitis or trichiasis or cataract* or hydrocephal* or cerebral palsy or muscular dystroph* or syndactyly* or radial club or amniotic band* or septic arthritis or osteomyelitis or flexor tenosynovitis or clubfoot or clubfeet or club-foot* or club-feet* or craniofacial* or cranio-facial* or frontoethmoidal meningoenceph* or hemorrhage* or hematoma* or spina bifida* or resuscitation* or schistosomias* or trachoma* or mediastinitis or buruli ulcer* or choledochal cyst* or cyst* echinococcosis or ilopsoas or epileps* or burr hole* or burn or burns or burned or scald* or burnt or thermal injur*).tw,kf. | 1624802 |
| 34 | (hypospadias* or epispadi* or cloaca* or cryptorchidism* or prolapse or phymosis or paraphymosis or hydrometrocolpos or (bladder adj2 exstroph*) or (undescen* adj2 test?s) or (buried adj1 penis) or (urinary adj2 (retention or lithiasis))).tw,kf. | 64934 |
| 35 | (hirschsprung* or ((megacolon or colon* or rectosigmoid or intestin*) adj4 (congenital or aganglion*)) or ((anal or anus or anorect* or rectal) adj3 (artificial* or malformation or anomal* or abnormal* or ectopic or stenosis or atres* or atroph* or imperforat* or inperforat* or praet* or pret*))).tw,kf. | 14047 |
| 36 | ((pierre-robin or apert*) adj2 (syndrome* or disease* or sequenc*).tw,kf. | 2407 |
| 37 | (cleft adj2 (lip* or palate*).tw,kf. | 24228 |
| 38 | (coarctation or (septal adj2 defect*) or (tetralogy adj2 fallot)).tw,kf. | 44183 |
| 39 | (brachial plexus adj2 (palsy or neuropath*).tw,kf. | 1564 |
| 40 | ((arthriti* or rheumat*) adj2 infect*).tw,kf. | 2853 |
| 41 | or/18-40 | 9782302 |
| 42 | exp Specialties, Surgical/ | 212232 |
| 43 | exp Surgical Procedures, Operative/ | 3401688 |
| 44 | su.fs. | 2150629 |
| 45 | (surger* or surgical* or operati* or reoperat* or bypass* or by-pass* or resect* or re-sect* or transplant* or procedure or procedures or debridement* or laparoscop* or laparotom*).tw,kf. | 4167558 |
| 46 | or/42-45 | 6158070 |
| 47 | 41 and 46 | 3046455 |
| 48 | 10 and 47 [Africa Peds Surg condition] | 10564 |
| 49 | (adenoidectom* or laryngectom* or laryngoplast* or laryngoscop* or pharygectom* or tonsillectom* or tympanoplast* or tracheostom* or tracheotom* or orchidopex* or orchiopex* or orchiectom* or orchidectom* or herniorrhaph* or hernioplast* or hernioplast* or herni*-plast* or herniotom* or circumcis* or gastrostom or ileostom* or colostom* or enterostom* or portoenterostom or roux-en-y or kasai or pyloromyotom* or piloromyotom* or pyloromiotom* or piloromiotom* or diverticulectom* or diverticulotom* or cholecystectom* or cholangiopancreatograph* or cholangio-pancreatograph* or choledoduodenostom* or choledo-duodenostom* or appendicectom* or appendectom* or splenectom* or pneumonectom* or amputation* or amputate* or craniotom* or craniostom* or hydrocelectom* or thoracostom* or fasciotom*).tw,kf. | 273533 |
| 50 | ((ophthalmolog* or eye* or vision or ocular or retina* or retinopath*) adj5 (operat* or procedur* or | 38688 |
|  | surger* or surgical*))).tw,kf. |  |
| 51 | ((perforation* or incision* or laceration*) adj3 (repair* or drain* or closure*))).tw,kf. | 9857 |
| 52 | ((tooth or teeth or dental or abcess) adj2 (extract* or drain*))).tw,kf. | 16161 |
| 53 | ((abdomen or abdominal or intestin* or bowel* or gastrointestin*) adj3 (ablat* or excis* or laparoscop* or laparotom* or operativ* or surger* or surgical* or reconstruct* or repair* or resect* or intraoperative* or perioperative* or perisurg* or postoperative* or postsurg* or preoperative* or presurg*))).tw,kf. | 76345 |
| 54 | (escharotom* or ((skin or derm*) adj2 (graft* or transplant*))).tw,kf. | 26742 |
| 55 | ((cancer or neoplas* or tumor* or tumour* or carcinom* or sarcoma*) adj3 (ablat* or excis* or laparoscop* or laparotom* or operativ* or surger* or surgical* or reconstruct* or repair* or resect* or biopsy or biopsie* or intraoperative* or perioperative* or perisurg* or postoperative* or postsurg* or preoperative* or presurg*))).tw,kf. | 224854 |
| 56 | (fundoplicat* or ((nissen* or toupet or dor) adj3 (operat* or procedur* or surger* or surgical*))).tw,kf. | 6810 |
| 57 | ((hernia* or extraperitoneal or preperitoneal or peritoneal or TEP or TAPP or umbilic* or inguinal or omphalocele* or exomphalos) adj3 (ablat* or excis* or laparoscop* or laparotom* or operativ* or surger* or surgical* or reconstruct* or repair* or resect* or intraoperative* or perioperative* or perisurg* or postoperative* or postsurg* or preoperative* or presurg*))).tw,kf. | 31190 |
| 58 | ((liver or hepatic or lung or lungs or pulmon* or kidney) adj3 (transplant* or graft*))).tw,kf. | 147121 |
| 59 | (thoroscop* or thoracotom* or pleurectom* or pleuroscop* or pleuracotom* or pleurotom* or (pleura* adj3 (endoscop* or incision*))).tw,kf. | 39993 |
| 60 | ((lung or lungs or pulmon* or wedge or trauma* or postrauma* or posttrauma* or neurotrauma* or fracture*) adj3 (ablat* or excis* or laparoscop* or laparotom* or operativ* or surger* or surgical* or reconstruct* or repair* or resect* or intraoperative* or perioperative* or perisurg* or postoperative* or postsurg* or preoperative* or presurg*))).tw,kf. | 110621 |
| 61 | or/49-60 | 916369 |
| 62 | 10 and 61 [Africa Peds Surg spec] | 4245 |
| 63 | Esophageal Atresia/ | 3855 |
| 64 | ((esophag* or oesophag* or endoesophag* or intraesophag* or tracheoesophag*) adj3 (atres* or atretic* or atroph*))).tw,kf. | 4852 |
| 65 | anus, imperforate/ | 2570 |
| 66 | ((anal or anus or anorect* or rectal) adj3 (artificial* or malformat* or mal-format* or anomal* or abnormal* or ectopic or stenosis or atres* or atroph* or imperforat* or inperforat* or praet* or pret* or fistula*))).tw,kf. | 10761 |
| 67 | Colorectal Surgery/ | 4040 |
| 68 | Rectal Diseases/ or exp *Rectal Diseases/ | 165790 |
| 69 | hirschsprung disease/ | 4788 |
| 70 | (hirschsprung* or ((megacolon or colon* or rectosigmoid or intestin*) adj3 (congenital* or aganglion*))).tw,kf. | 7050 |
| 71 | Hernias, Diaphragmatic, Congenital/ | 5231 |
| 72 | (agene* adj2 (hemidiaphragm* or diaphragm* or ((unilat* or hern*) adj1 diaphragm*))).tw,kf. | 86 |
| 73 | ((bochdalek* or morgagni*) adj2 (hernia* or defect*))).tw,kf. | 1310 |
| 74 | ((congenital* or neonat* or neo-nat* or newborn* or new-born* or birth* or maternal* or fetal or fetus* or fetu or feto or foet* or prenatal* or pre-natal* or antenatal* or ante-natal* or trimester* or pregnan* or uter* or preterm* or pre-term* or premature* or pre-mature* or preemie*) adj5 (posterolateral* or substernal*) adj2 hernia*))).tw,kf. | 78 |
| 75 | ((congenital* or neonat* or neo-nat* or newborn* or new-born* or birth* or maternal* or fetal or | 5591 |
|  | fetus* or fetu or feto or foet* or prenatal* or pre-natal* or antenatal* or ante-natal* or trimester* or pregnan* or uter* or preterm* or pre-term* or premature* or pre-mature* or premie*) adj2 diaphragm* adj2 (hernia* or defect*)).tw,kf. |  |
| 76 | (congenital* and hernia* and diaphragm*).tw,kf. | 6135 |
| 77 | musculoskeletal abnormalities/ | 1595 |
| 78 | funnel chest/ | 2596 |
| 79 | pectus carinatum/ | 133 |
| 80 | ((pectus or chest) adj1 (funnel or sunken or excavatum or carinatum)).tw,kf. | 3263 |
| 81 | or/63-80 [Specific surg cond] | 200581 |
| 82 | 10 and 81 [Africa Peds Specific Surg] | 594 |
| 83 | 17 or 48 or 62 or 82 [Africa Peds Surg] | 23054 |
| 84 | Attitude of Health Personnel/ | 129230 |
| 85 | exp Health Personnel/ | 576457 |
| 86 | exp Administrative Personnel/ | 41817 |
| 87 | Social Workers/ | 918 |
| 88 | or/84-87 | 653451 |
| 89 | Stereotyping/ | 11703 |
| 90 | Social Stigma/ | 11205 |
| 91 | Prejudice/ | 25336 |
| 92 | Social Discrimination/ | 1517 |
| 93 | or/89-92 | 45347 |
| 94 | 88 and 93 | 8245 |
| 95 | ((attitude* or bias* or perception* or antipathy or stigma* or anti-stigma* or stereotyp* or prejudice* or discriminat* or bias* or microaggression* or micro-aggression* or negative attitude*) adj5 (((health* or hospital* or clinic* or medic*) adj2 (professional* or personnel* or staff* or employee* or provider*)) or doctor* or physician* or surgeon* or resident* or nurse or nurses or allied health or social worker* or secretar* or admin* agent*)).tw,kf. | 39729 |
| 96 | exp Health Services Accessibility/ | 121610 |
| 97 | ((health* or medic* or surg* or doctor* or clinic* or appointment*) adj3 (access* or equit* or inequit* or inaccess* or barrier* or burden*)).tw,kf. or ((health* or medic* or surg* or doctor* or clinic* or appointment*) adj5 (access* or equit* or inequit* or inaccess* or barrier* or delay* or distance* or wait* or long-term or complication* or factor* or influenc*)).ti,kf. | 215550 |
| 98 | Sex/ | 7712 |
| 99 | Sex Characteristics/ | 59268 |
| 100 | "Sex Factors"/ | 277777 |
| 101 | exp "sexual and gender minorities"/ | 12037 |
| 102 | Gender Equity/ | 387 |
| 103 | Sexism/ | 2928 |
| 104 | exp Gender Identity/ | 22093 |
| 105 | Intersex Persons/ | 13 |
| 106 | Transgender Persons/ | 5188 |
| 107 | (LGBT* or GLBT* or LGB or LGBQ* or LGBS* or M2F or GLB or GLBQ* or GLBs* or 2LGBTQS* or heteroflexible* or heterosex* or cisgend* or GBMSM* or msm or TGNC* or YTW or | 389643 |
|  | gay or gays or lesbi* or bisexu#l* or transgender* or bicurious* or transpeople* or asexual* or transvestite or cross sex* or crosssex* or crossgender* or nonbinary or non-binary or F2M or transperson* or transsexual* or homosexual* or intersex* or queer* or two-spirit* or same-sex or male-to-female or boy-to-girl or (trans adj2 (people or individual or individuals or person or persons or sexual* or male or female or youth* or population* or gender)) or ((gender* or sex or sexual) adj2 (change or dysphoria or reversal or identit* or reassign* or transform* or transition* or minorit* or sexual* or binary or disaggreg*))).tw,kf. |  |
| 108 | (gender* or girl* or boy*).ti,kf. | 116060 |
| 109 | (gender* or sex or female* or male* or wom?n or m?n or girl* or boy* or parent? or guardian* or child* or caregiver*).ti,kf. or (gender* or sex or female* or male* or wom?n or m?n or girl* or boy* or parent? or guardian* or child* or caregiver*).ab. /freq=3 | 2873690 |
| 110 | (access* or equit* or inequit* or inaccess* or barrier* or delay* or distance* or wait* or long-term or complication* or burden* or factor* or influenc* or time or location*).tw,kf. and (surger* or surgical* or operati* or reoperat* or bypass* or by-pass* or resect* or re-sect* or transplant* or procedure or procedures or debridement* or laparoscop* or laparotom*).ti,kf. | 626929 |
| 111 | 109 and 110 | 62369 |
| 112 | (or/94-108) or 111 | 1151078 |
| 113 | 83 and 112 [Africa Peds Surg & Gender] | 4877 |
| 114 | remove duplicates from 113 | 4873 |

#### Web of Science [Clarivate Analytics] (March 31, 2022)

|  |  |  |
| --- | --- | --- |
| 38 | #36 NOT #37 | 411 |
| 37 | PMID=(0* or 1* OR 2* OR 3* OR 4* OR 5* OR 6* OR 7* OR 8* OR 9*) | 25,763,788 |
| 36 | #35 AND #27 | 2,270 |
| 35 | #34 OR #31 OR #30 OR #29 OR #28 | 1,540,147 |
| 34 | #33 AND #32 | 30,099 |
| 33 | TI=((access* or equit* or inequit* or inaccess* or barrier* or delay* or distance* or wait* or long-term or complication* or burden* or factor* or influenc* or time or location*) and (surger* or surgical* or operati* or reoperat* or bypass* or by-pass* or resect* or re-sect* or transplant* or procedure or procedures or debridement* or laparoscop* or laparotom*)) | 172,798 |
| 32 | TI=(gender* or sex or female* or male* or wom?n or m?n or girl* or boy* or parent? or guardian* or child* or caregiver*) or AB=(gender* or sex or female* or male* or wom?n or m?n or girl* or boy* or parent? or guardian* or child* or caregiver*) or AB=(gender* or sex or female* or male* or wom?n or m?n or girl* or boy* or parent? or guardian* or child* or caregiver*) | 7,183,014 |
| 31 | TI=(gender* or girl* or boy*) | 278,259 |
| 30 | TI=(LGBT* or GLBT* or LGB or LGBQ* or LGBS* or M2F or GLB or GLBQ* or GLBs* or 2LGBTQS* or heteroflexible* or heterosex* or cisgend* or GBMSM* or msm or TGNC* or YTW or gay or gays or lesbi* or bisexual* or transgender* or bicurious* or transpeople* or asexual* or transvestite or cross sex* or crosssex* or crossgender* or nonbinary or non-binary or F2M or transperson* or transsexual* or homosexual* or intersex* or queer* or two-spirit* or same-sex or male-to-female or boy-to-girl or (trans NEAR/2 (people or individual or individuals or person or persons or sexual* or male or female or youth* or population* or gender)) or ((gender* or sex or sexual) NEAR/2 (change or dysphoria or reversal or identit* or reassign* or transform* or transition* or minorit* or sexual* or binary or disaggreg*))) OR AB=(LGBT* or GLBT* or LGB or LGBQ* or LGBS* or M2F or GLB or GLBQ* or GLBs* or 2LGBTQS* or heteroflexible* or heterosex* or cisgend* or GBMSM* or msm or TGNC* or YTW or gay or gays or lesbi* or bisexual* or transgender* or bicurious* or transpeople* or asexual* or transvestite or cross sex* or crosssex* or crossgender* or nonbinary or non-binary or F2M or transperson* or transsexual* or homosexual* or intersex* or queer* or two-spirit* or same-sex or male-to-female or boy-to-girl or (trans NEAR/2 (people or individual | 509,324 |
|  | or individuals or person or persons or sexual* or male or female or youth* or population* or gender)) or ((gender* or sex or sexual) NEAR/2 (change or dysphoria or reversal or identit* or reassign* or transform* or transition* or minorit* or sexual* or binary or disaggreg*)) |  |
| 29 | TI=((health* or medic* or surg* or doctor* or clinic* or appointment*) NEAR/3 (access* or equit* or inequit* or inaccess* or barrier* or burden*)) or TI=((health* or medic* or surg* or doctor* or clinic* or appointment*) NEAR/3 (access* or equit* or inequit* or inaccess* or barrier* or burden*)) or AB=((health* or medic* or surg* or doctor* or clinic* or appointment*) NEAR/5 (access* or equit* or inequit* or inaccess* or barrier* or delay* or distance* or wait* or "long-term" or complication* or factor* or influenc*)) | 656,068 |
| 28 | TI=((attitude* or bias* or perception* or antipathy or stigma* or anti-stigma* or stereotyp* or prejudice* or discriminat* or bias* or microaggression* or micro-aggression* or negative attitude*) AND (((health* or hospital* or clinic* or medic*) NEAR/2 (professional* or personnel* or staff* or employee* or provider*)) or doctor* or physician* or surgeon* or resident* or nurse or nurses or allied health or social worker* or secretar* or admin* agent*)) OR AB=((attitude* or bias* or perception* or antipathy or stigma* or anti-stigma* or stereotyp* or prejudice* or discriminat* or bias* or microaggression* or micro-aggression* or negative attitude*) AND (((health* or hospital* or clinic* or medic*) NEAR/2 (professional* or personnel* or staff* or employee* or provider*)) or doctor* or physician* or surgeon* or resident* or nurse or nurses or allied health or social worker* or secretar* or admin* agent*)) | 159,408 |
| 27 | #26 AND #5 | 10,936 |
| 26 | #25 OR #24 OR #23 OR #22 OR #21 OR #20 OR #19 OR #18 OR #17 OR #16 OR #12 OR #11 OR #13 OR #14 OR #15 OR #10 OR #9 OR #8 OR #7 OR #6 | 6,586,371 |
| 25 | TI=((pectus or chest) NEAR/1 (funnel or sunken or excavatum or carinatum)) OR AB=((pectus or chest) NEAR/1 (funnel or sunken or excavatum or carinatum)) | 2,602 |
| 24 | TI=((congenital* and hernia* and diaphragm*) OR AB=((congenital* and hernia* and diaphragm*)) | 5,754 |
| 23 | TI=((congenital* or neonat* or neo-nat* or newborn* or new-born* or birth* or maternal* or fetal or fetus* or fetu or feto or foet* or prenatal* or pre-natal* or antenatal* or ante-natal* or trimester* or pregnan* or uter* or preterm* or pre-term* or premature* or pre-mature* or preemie*) NEAR/2 diaphragm* NEAR/2 (hernia* or defect*)) OR AB=((congenital* or neonat* or neo-nat* or newborn* or new-born* or birth* or maternal* or fetal or fetus* or fetu or feto or foet* or prenatal* or pre-natal* or antenatal* or ante-natal* or trimester* or pregnan* or uter* or preterm* or pre-term* or premature* or pre-mature* or preemie*) NEAR/2 diaphragm* NEAR/2 (hernia* or defect*)) | 5,598 |
| 22 | TI=((congenital* or neonat* or neo-nat* or newborn* or new-born* or birth* or maternal* or fetal or fetus* or fetu or feto or foet* or prenatal* or pre-natal* or antenatal* or ante-natal* or trimester* or pregnan* or uter* or preterm* or pre-term* or premature* or pre-mature* or preemie*) NEAR/5 (posterolateral* or substernal*) NEAR/2 hernia*) OR AB=((congenital* or neonat* or neo-nat* or newborn* or new-born* or birth* or maternal* or fetal or fetus* or fetu or feto or foet* or prenatal* or pre-natal* or antenatal* or ante-natal* or trimester* or pregnan* or uter* or preterm* or pre-term* or premature* or pre-mature* or preemie*) NEAR/5 (posterolateral* or substernal*) NEAR/2 hernia*) | 64 |
| 21 | TI=((bochdalek* or morgagni*) NEAR/2 (hernia* or defect*)) OR AB=((bochdalek* or morgagni*) NEAR/2 (hernia* or defect*)) | 1,052 |
| 20 | TI=(agene* NEAR/2 (hemidiaphragm* or diaphragm* or ((unilat* or hern*) NEAR/1 diaphragm*))) OR AB=(agene* NEAR/2 (hemidiaphragm* or diaphragm* or ((unilat* or hern*) NEAR/1 diaphragm*))) | 120 |
| 19 | TI=(hirschsprung* or ((megacolon or colon* or rectosigmoid or intestin*) NEAR/3 (congenital* or aganglion*))) OR AB=(hirschsprung* or ((megacolon or colon* or rectosigmoid or intestin*) NEAR/3 (congenital* or aganglion*))) | 6,519 |
| 18 | TI=((anal or anus or anorect* or rectal) NEAR/3 (artificial* or malformat* or mal-format* or anomal* or abnormal* or ectopic or stenosis or atres* or atroph* or imperforat* or imperforat* or praet* or pret* or fistula*)) OR AB=((anal or anus or anorect* or rectal) NEAR/3 (artificial* or malformat* or mal-format* or anomal* or abnormal* or ectopic or stenosis or atres* or atroph* or imperforat* or imperforat* or praet* or pret* or fistula*)) | 9,279 |
| 17 | TI=((lung or lungs or pulmon* or wedge or trauma* or postrauma* or posttrauma* or | 109,572 |
|  | neurotrauma* or fracture*) NEAR/3 (ablat* or excis* or laparoscop* or laparotom* or operativ* or surger* or surgical* or reconstruct* or repair* or resect* or intraoperative* or perioperative* or perisurg* or postoperative* or postsurg* or preoperative* or presurg*)) OR AB=((lung or lungs or pulmon* or wedge or trauma* or postrauma* or postrauma* or neurotrauma* or fracture*) NEAR/3 (ablat* or excis* or laparoscop* or laparotom* or operativ* or surger* or surgical* or reconstruct* or repair* or resect* or intraoperative* or perioperative* or perisurg* or postoperative* or postsurg* or preoperative* or presurg*)) |  |
| 16 | TI=(thoroscop* or thoracotom* or pleurectom* or pleuroscop* or pleuracotom* or pleurotom* or (pleura* NEAR/3 (endoscop* or incision*))) OR AB=(thoroscop* or thoracotom* or pleurectom* or pleuroscop* or pleuracotom* or pleurotom* or (pleura* NEAR/3 (endoscop* or incision*))) | 31,645 |
| 15 | TI=((liver or hepatic or lung or lungs or pulmon* or kidney) NEAR/3 (transplant* or graft*)) OR AB=((liver or hepatic or lung or lungs or pulmon* or kidney) NEAR/3 (transplant* or graft*)) | 198,481 |
| 14 | TI=((hernia* or extraperitoneal or preperitoneal or peritoneal or TEP or TAPP or umbilic* or inguinal or omphalocele* or exomphalos) NEAR/3 (ablat* or excis* or laparoscop* or laparotom* or operativ* or surger* or surgical* or reconstruct* or repair* or resect* or intraoperative* or perioperative* or perisurg* or postoperative* or postsurg* or preoperative* or presurg*)) OR AB=((hernia* or extraperitoneal or preperitoneal or peritoneal or TEP or TAPP or umbilic* or inguinal or omphalocele* or exomphalos) NEAR/3 (ablat* or excis* or laparoscop* or laparotom* or operativ* or surger* or surgical* or reconstruct* or repair* or resect* or intraoperative* or perioperative* or perisurg* or postoperative* or postsurg* or preoperative* or presurg*)) | 33,115 |
| 13 | TI=(fundoplicat* or ((nissen* or toupet or dor) NEAR/3 (operat* or procedur* or surger* or surgical*))) OR AB=(fundoplicat* or ((nissen* or toupet or dor) NEAR/3 (operat* or procedur* or surger* or surgical*))) | 6,796 |
| 12 | TI=((cancer or neoplas* or tumor* or tumour* or carcinom* or sarcoma*) NEAR/3 (ablat* or excis* or laparoscop* or laparotom* or operativ* or surger* or surgical* or reconstruct* or repair* or resect* or biopsy or biopsie* or intraoperative* or perioperative* or perisurg* or postoperative* or postsurg* or preoperative* or presurg*)) OR AB=((cancer or neoplas* or tumor* or tumour* or carcinom* or sarcoma*) NEAR/3 (ablat* or excis* or laparoscop* or laparotom* or operativ* or surger* or surgical* or reconstruct* or repair* or resect* or biopsy or biopsie* or intraoperative* or perioperative* or perisurg* or postoperative* or postsurg* or preoperative* or presurg*)) | 270,089 |
| 11 | TI=(escharotom* or ((skin or derm*) NEAR/2 (graft* or transplant*))) OR AB=(escharotom* or ((skin or derm*) NEAR/2 (graft* or transplant*))) | 19,519 |
| 10 | TI=((abdomen or abdominal or intestin* or bowel* or gastrointestin*) NEAR/3 (ablat* or excis* or laparoscop* or laparotom* or operativ* or surger* or surgical* or reconstruct* or repair* or resect* or intraoperative* or perioperative* or perisurg* or postoperative* or postsurg* or preoperative* or presurg*)) OR AB=((abdomen or abdominal or intestin* or bowel* or gastrointestin*) NEAR/3 (ablat* or excis* or laparoscop* or laparotom* or operativ* or surger* or surgical* or reconstruct* or repair* or resect* or intraoperative* or perioperative* or perisurg* or postoperative* or postsurg* or preoperative* or presurg*)) | 77,083 |
| 9 | TI=((tooth or teeth or dental or abcess) NEAR/2 (extract* or drain*)) OR AB=((tooth or teeth or dental or abcess) NEAR/2 (extract* or drain*)) | 14,121 |
| 8 | TI=((ophthalmolog* or eye* or vision or ocular or retina* or retinopath*) NEAR/5 (operat* or procedur* or surger* or surgical*)) OR AB=((ophthalmolog* or eye* or vision or ocular or retina* or retinopath*) NEAR/5 (operat* or procedur* or surger* or surgical*)) | 39,064 |
| 7 | TI=(adenoidectom* or laryngectom* or laryngoplast* or laryngoscop* or pharygectom* or tonsillectom* or tympanoplast* or tracheostom* or tracheotom* or orchidopex* or orchiopex* or orchiectom* or orchidectom* or herniorrhaph* or hernioplast* or hernioplast* or herni*-plast* or herniotom* or circumcis* or gastrostom or ileostom* or colostom* or enterostom* or portoenterostom or roux-en-y or kasai or pyloromyotom* or piloromyotom* or pyloromiotom* or piloromiotom* or diverticulectom* or diverticulotom* or cholecystectom* or cholangiopancreatograph* or cholangio-pancreatograph* or choledoduodenostom* or choledo-duodenostom* or appendicectom* or appendectom* or splenectom* or pneumonectom* or amputation* or amputate* or craniotom* or craniostom*) | 224,503 |
|  | or hydrocelectom* or thoracostom* or fasciotom*) or AB=(adenoidectom* or laryngectom* or laryngoplast* or laryngoscop* or pharygectom* or tonsillectom* or tympanoplast* or tracheostom* or tracheotom* or orchidopex* or orchiopex* or orchiectom* or orchidectom* or herniorrhaph* or hernioplast* or hernioplast* or herni*.plast* or herniotom* or circumcis* or gastrostom or ileostom* or colostom* or enterostom* or portoenterostom or roux-en-y or kasai or pyloromyotom* or piloromyotom* or pyloromiotom* or piloromiotom* or diverticulectom* or diverticulotom* or cholecystectom* or cholangiopancreatograph* or cholangio-pancreatograph* or choledoduodenostom* or choledo-duodenostom* or appendicectom* or appendectom* or splenectom* or pneumonectom* or amputation* or amputate* or craniotom* or craniostom* or hydrocelectom* or thoracostom* or fasciotom*) |  |
| 6 | TI=(surger* or surgic* or surgeon* or operati* or reoperat* or bypass* or by-pass* or resect* or re-sect* or transplant* or procedure* or debridement* or laparoscop* or laparotom* or postop*) OR AB=(surger* or surgic* or surgeon* or operati* or reoperat* or bypass* or by-pass* or resect* or re-sect* or transplant* or procedure* or debridement* or laparoscop* or laparotom* or postop*) | 6,388,614 |
| 5 | #3 AND #4 | 160,663 |
| 4 | TS=(newborn* or new-born* or neonat* or neo-nat* or infan* or child* or paediatr* or pediater* or baby* or babies* or toddler* or kid or kids or boy* or girl* or adolesc* or juvenile* or teen* or youth* or pubescen* or preadolesc* or prepubesc* or preteen*) | 3,732,184 |
| 3 | #1 OR #2 | 1,376,193 |
| 2 | TS=((Porto-Novo or Hogbonou or Cotonou or Kutonu or Ouagadougou or Ouaga or Bujumbura or Usumbura or Bangui or Bangi or N'Djamena or Ndjamenia or Fort Lamy or Moroni or Kinshasa or Asmara or Asmera or Addis Ababa or Addis Abeba or Banjul or Bathurst or Conakry or Bissau or Port-au-Prince or Monrovia or Antananarivo or Tananarive or Tana or Lilongwe or Bamako or Maputo or Lourenco Marques or Niamey or Kigali or Freetown or Free-town or Mogadishu or Xamar or Hamar or Muqdisho or Maqadishu or Juba or Dodoma or Dar es Salaam or Lome or Kampala or Harare or Salisbury or Thimphu or Thimbu or La Paz or Praia or Yaounde or Jaunde or Brazzaville or Yamoussoukro or Cairo or Accra or Nairobi or Frunze or Maseru or Nouakchott or Rabat or Managua or Abuja or Lagos or Port Moresby or Moresby or Khartoum or Mbabane or Embabane or Lobamba or sanada or Lusaka or Luanda)) | 64,720 |
| 1 | TS=(Abuja or Abyssinia or Accra or Addis Ababa or Addis Abeba or Africa* or Algeria or Algiers or Angola or Antananarivo or Asmara or Asmera or Bamako or Bangi or Bangui or Banjul or Basutoland or Benin or Bissau or Bloemfontein or Botswana or Brazzaville or Bujumbura or Burundi or Burkina Faso or Burundi or Cabo Verde or Cairo or Cameroon or Cameroun or Cape Town or Cape Verde or Cap-Vert or Centrafrican or Centrafrique or Chad or Comores or Comoro or Comoros or Conakry or Congo or Cote Diivoire or Cote D'ivoire or Cotonou or Dahomey or Dakar or Dar Es Salaam or Djibouti or Dodoma or Egypt* or Embabane or Eritrea or Ethiopia or Eswatini or Freetown or Free-Town or Gabon or Gaborone or Gambia or Ghana or Gold Coast or Guinea or Harare or Ifni or Ivory Coast or Jaunde or Juba or Kampala or Katanga or Kenya or Khartoum or Kigali or Kinshasa or Kutonu or Leopoldville or Lesotho or Liberia or Libreville or Libya or Lilongwe or Lobamba or Lome or Luanda or Lusaka or Madagascar or Malabo or Malagasy or Malawi or Mali or Maputo or Maqadishu or Maseru or Mauretania or Mauritania or Mauritius or Mayotte or Mbabane or Mocambique or Mogadishu or Monrovia or Morocco or Moroni or Mozambic or Mozambique or Muqdisho or Nairobi or Namibia or Ndjamenia or N'Djamena or Niamey or Niger or Nigeria or Nouakchott or Nyasaland or Ouaga or Ouagadougou or Oyala or Port Louis or Porto-Novo or Praia or Pretoria or Rabat or Rhodesia or Ruanda or Rwanda or Sao Tome or Saharan* or Senegal* or Senegambia or Seychelles or Sierra Leone or Somali* or Subsahar* or Sudan or Swaziland or Tana or Tananarive or Tanganyika or Tanzania or Tchad or Togo or Togoland or Togolese or Tripoli or Tshad or Tunis or Tunisia or Ubangi-Shari or Uganda or "United Arab Republic" or Upper Volta or Urundi or Usumbura or Windhoek or Yamoussoukro or Yaounde or Zaire or Zambia or Zanzibar or Zimbabwe) | 1,368,940 |

## Appendix B PRISMA Checklist

| Section / Topic | Item No. | Checklist item | Reported on Page No. |
| --- | --- | --- | --- |

| TITLE |  |  |  |
| --- | --- | --- | --- |
| Title | 1 | Identify the report as a systematic review, meta-analysis, or both. | 2 |
| ABSTRACT |  |  |  |
| Structured Summary | 2 | Provide a structured summary including, as applicable: background; objectives; data sources; study eligibility criteria, participants, and interventions; study appraisal and synthesis methods; results; limitations; conclusions and implications of key findings;systematic review registration number. |  |
|  |  |  | 2-8 |
| INTRODUCTION |  |  |  |
| Rationale | 3 | Describe the rationale for the review in the context of what is already known. | 2,3 |
| Objectives | 4 | Provide an explicit statement of questions being addressed with reference to participants, interventions, comparisons, outcomes, and study design (PICOS). |  |
|  |  |  | 2,3 |
| METHODS |  |  |  |
| Protocol and registration | 5 | Indicate if a review protocol exists, if and where it can be accessed (e.g., Web address), and, if available, provide registration information including registration number. |  |
|  |  |  | 4 |
| Eligibility criteria | 6 | Specify study characteristics (e.g., PICOS, length of follow-up) and report characteristics (e.g., years considered, language, publication status) used as criteria for eligibility, giving rationale. |  |
|  |  |  | 3-5 |
| Information sources | 7 | Describe all information sources (e.g., databases with dates of coverage, contact with study authors to identify additional studies) in the search and date last searched. |  |
|  |  |  | 4 |
| Search | 8 | Present full electronic search strategy for at least one database, including any |  |
|  |  | limits used, such that it could be repeated. | 4, Appendix A |
| Study selection | 9 | State the process for selecting studies (i.e., screening, eligibility, included in systematic review, and, if applicable, included in the meta-analysis). | 4,5 |
| Data collection process | 10 | Describe method of data extraction from reports (e.g., piloted forms, independently, in duplicate) and any processes for obtaining and confirming data from investigators. | 4,5 |
| Data items | 11 | List and define all variables for which data were sought (e.g., PICOS, funding sources) and any assumptions and simplifications made. | 3-5 |
| Risk of bias in individual studies | 12 | Describe methods used for assessing risk of bias of individual studies (including specification of whether this was done at the study or outcome level), and how this information is to be used in any data synthesis. | 5,7 |

| Section / Topic | Item No. | Checklist item | Reported on Page No. |
| --- | --- | --- | --- |
| Summary Measures | 13 | State the principal summary measures (e.g., risk ratio, difference in means). | 5-7 |
| Synthesis of results | 14 | Describe the methods of handling data and combining results of studies, if done, including measures of consistency (e.g., $I^2$) for each meta-analysis. | 5-7 |
| Risk of bias across studies | 1<br>5 | Specify any assessment of risk of bias that may affect the cumulative evidence (e.g., publication bias, selective reporting within studies). |  |
|  |  |  | 7 |
| Additional Analyses | 1<br>6 | Describe methods of additional analyses (e.g., sensitivity or subgroup analyses, meta-regression), if done, indicating which were pre-specified. |  |
|  |  |  | 6,7 |
| RESULTS |  |  |  |
| Study selection | 1<br>7 | Give numbers of studies screened, assessed for eligibility, and included in the review, with reasons for exclusions at each stage, ideally with a flow diagram. |  |
|  |  |  | 5,6,12 |
| Study Characteristics | 1<br>8 | For each study, present characteristics for which data were extracted (e.g., study size, PICOS, follow-up period) and provide the citations. |  |
|  |  |  | 6, 13 |
| Risk of bias within studies | 1<br>9 | Present data on risk of bias of each study and, if available, any outcome level assessment (see item 12). |  |
|  |  |  | 7, 18 |
| Results of individual studies | 2<br>0 | For all outcomes considered (benefits or harms), present, for each study: (a) simple summary data for each intervention group<br>(b) effect estimates and confidence intervals, ideally with a forest plot. |  |
|  |  |  | 6,7,16,17 |
| Synthesis of results | 2<br>1 | Present results of each meta-analysis done, including confidence intervals and measures of consistency. | 6,7,16,17 |
| Risk of bias across studies | 2 | Present results of any assessment of risk of bias across studies (see Item 15). |  |
|  | 2 |  | 7, 18 |
| Additional analysis | 2 | Give results of additional analyses, if done (e.g., sensitivity or subgroup analyses, meta-regression [see Item 16]). |  |
|  | 3 |  | 7 |
| DISCUSSION |  |  |  |
| Summary of evidence | 2 | Summarize the main findings including the strength of evidence for each main outcome; consider their relevance to key groups (e.g., healthcare providers, users, and policy makers). |  |
|  | 4 |  | 7,8 |
| Limitations | 2 | Discuss limitations at study and outcome level (e.g., risk of bias), and at review-level (e.g., incomplete retrieval of identified research, reporting bias). |  |
|  | 5 |  | 7 |
| Conclusions | 2 | Provide a general interpretation of the results in the context of other evidence, and implications for future research. |  |
|  | 6 |  | 7,8 |
| FUNDING |  |  |  |

| Section/Topic | Item No. | Checklist item | Reported on Page No. |
| --- | --- | --- | --- |
| Funding | 27 | Describe sources of funding for the systematic |  |
|  |  | review and other support (e.g., supply of data); role of funders for the systematic review. | 8 |

*From: Moher D, Liberati A, Tetzlaff J, Altman DG, The PRISMA Group (2009). Preferred Reporting Items for Systematic Reviews and Meta-Analyses: The PRISMA Statement. PLoS Med 6(6): e1000097. doi:10.1371/journal.pmed1000097*

### PRISMA-S Checklist

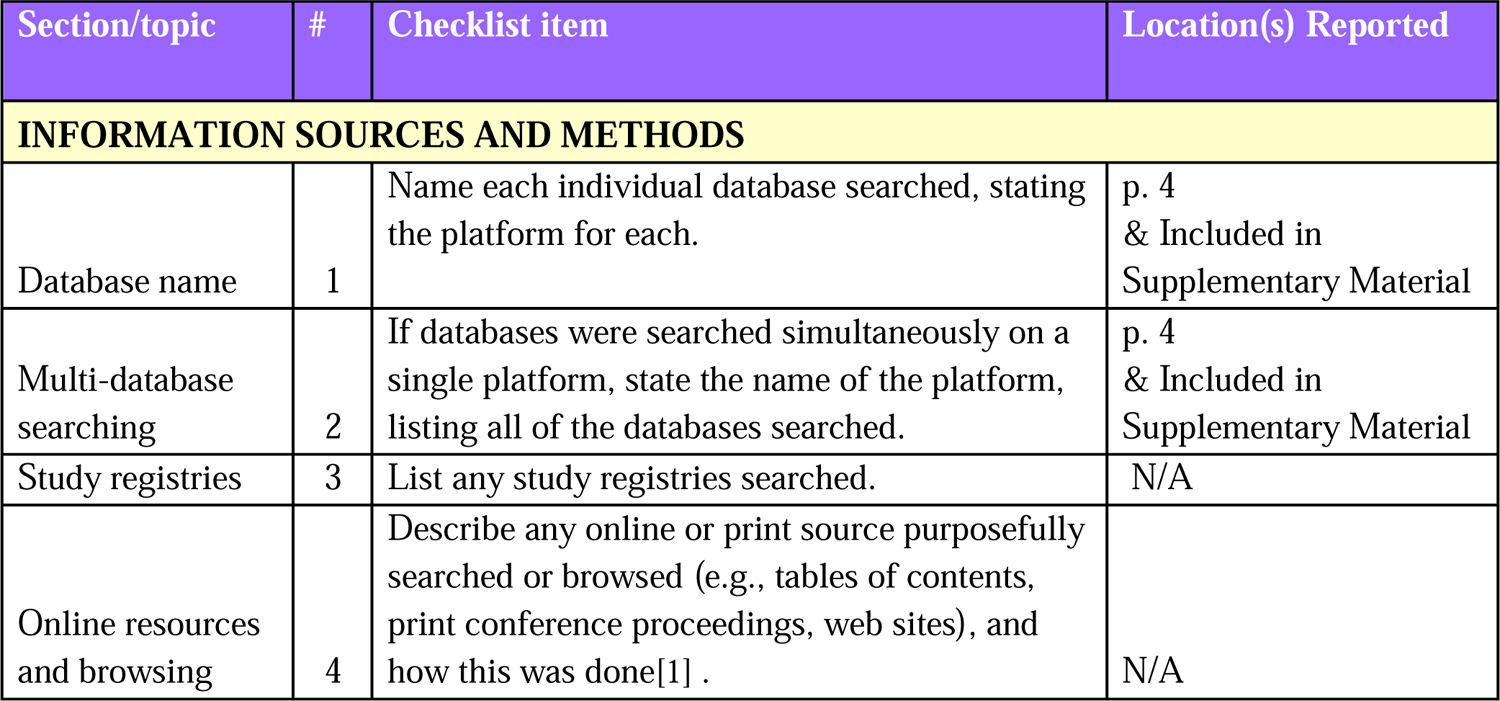
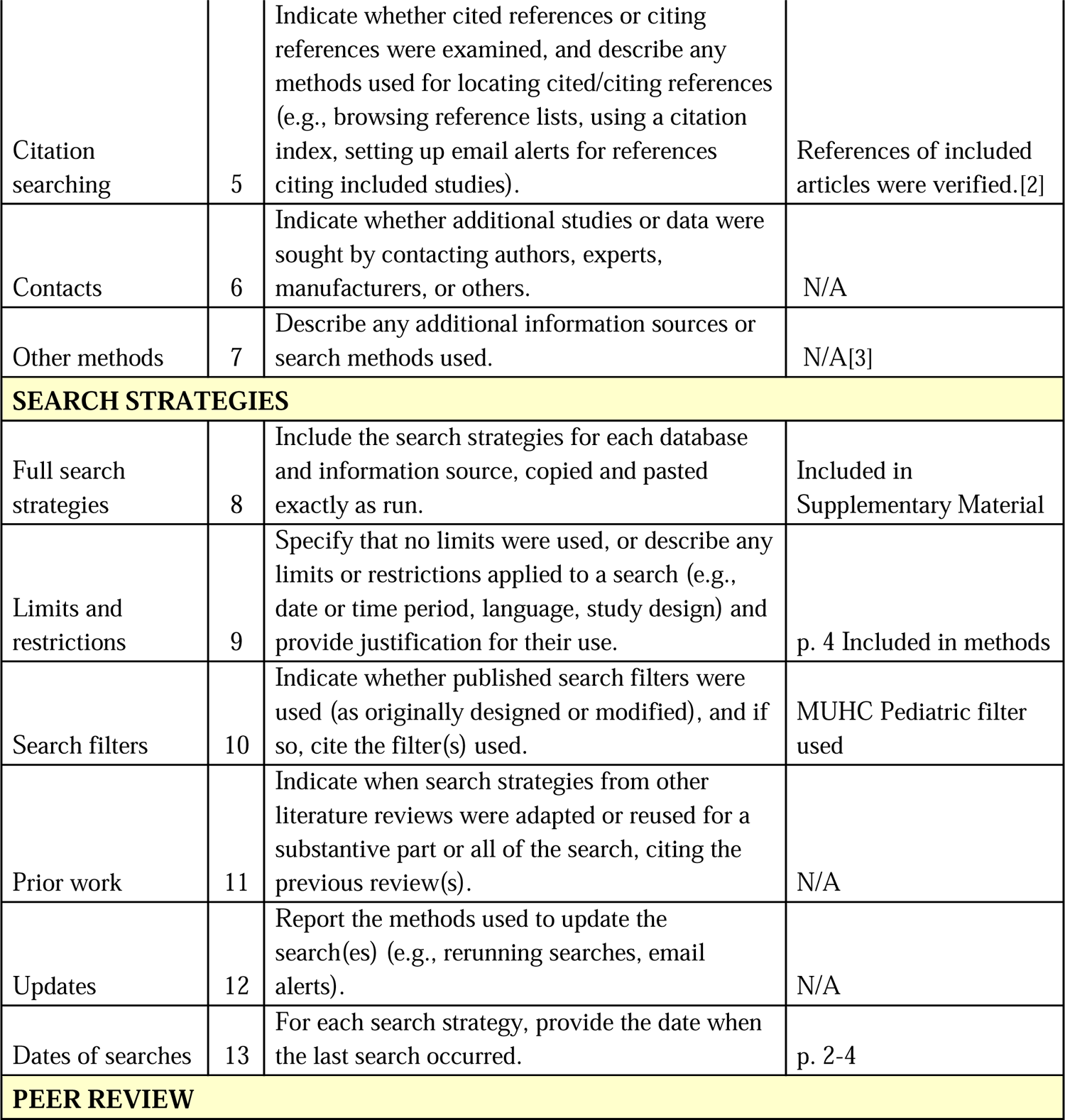
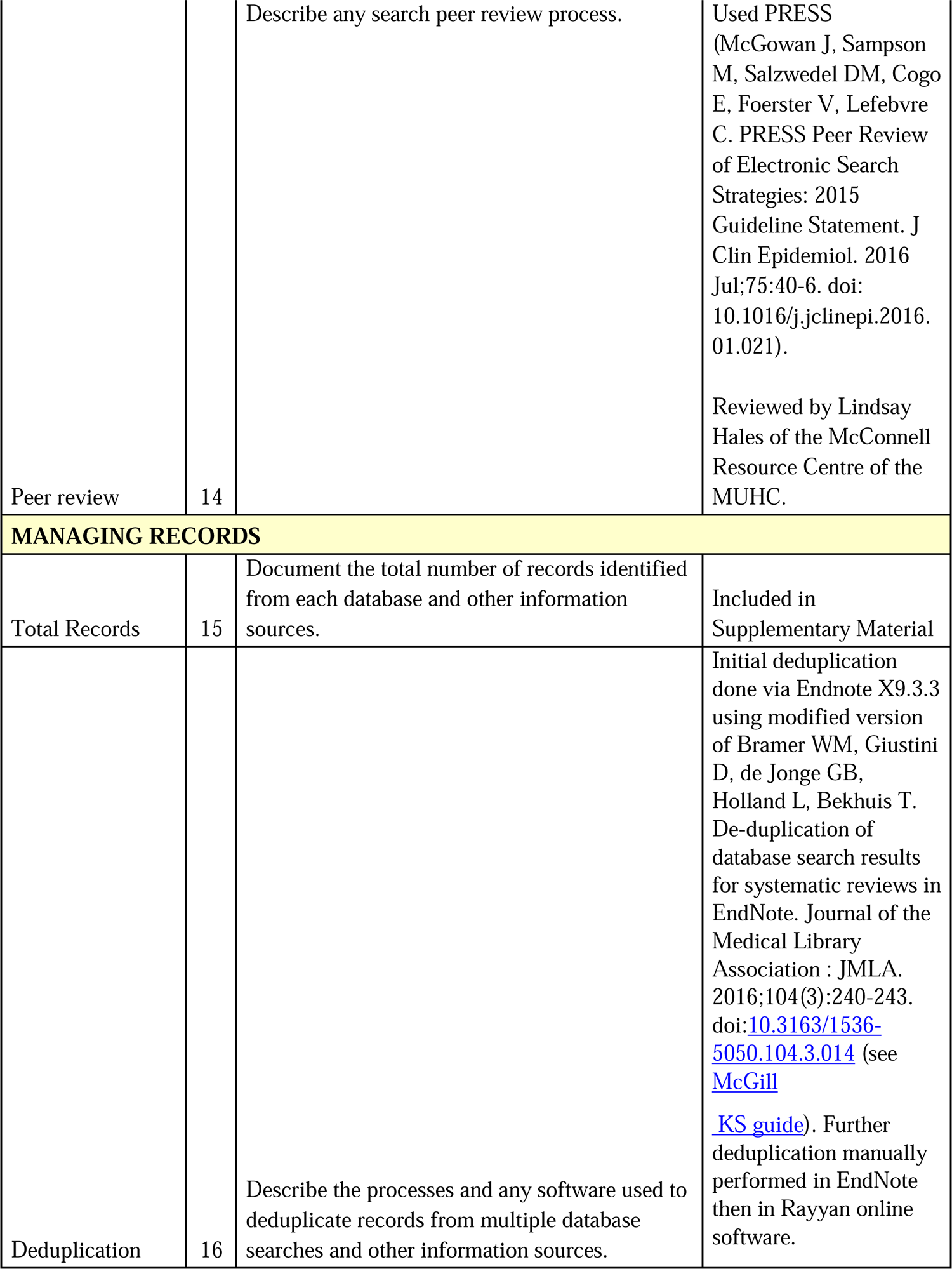

PRISMA-S: An Extension to the PRISMA Statement for Reporting Literature Searches in Systematic Reviews, Rethlefsen ML, Kirtley S, Waffenschmidt S, Ayala AP, Moher D, Page MJ, Koffel JB, PRISMA-S Group. Last updated February 27, 2020.

## Appendix C Access, adverse postoperative outcomes, and mortality calculations

| **Terms** | **Calculations** |
| --- | --- |
| Proportion of deaths by same sex | Proportion of deaths / total number of same sex cohort |
| Proportion of deaths by total deaths | Proportion of deaths / total number of deaths |
| Mortality Rate | Proportion of deaths / total number in cohort |
| Sex ratio for surgical access | Number of males in cohort / number of females in cohort |
| Sex ratio of mortality | Male mortality / female mortality |
| Sex ratio of the proportion of deaths by sex | [(Proportion of male deaths / total number of males in cohort) / (proportion of female deaths / total number of females in cohort)] |
| Sex ratio of complications excluding death | Male complications / female complications |

## Appendix D Finalized list of studies for meta-analysis (n = 54)

| **Study No** | **Authors** | **Year** | **Country** | **Study Design** | **Cohort Size** | **Males** | **Females** | **Surgical Specialty** |
| --- | --- | --- | --- | --- | --- | --- | --- | --- |
| 1 | Wubie AB et al. [1*] | 2022 | Ethiopia | Retrospective | 337 | 197 | 140 | Neurosurgery |
| 2 | Ullrich SJ et al. [2*] | 2022 | Uganda | Prospective | 3194 | 1941 | 1253 | Pediatric surgery |
| 3 | Davis JR et al. [3] | 2022 | Rwanda | Retrospective | 112 | 58 | 54 | Pediatric surgery |
| 4 | Ajao AE and Adeniran JO [4*] | 2022 | Nigeria | Cross-sectional | 336 | 229 | 107 | Pediatric surgery |
| 5 | Zungu T et al. [5*] | 2021 | Malawi | Prospective | 102 | 75 | 27 | Ophthalmology |
| 6 | Swart O et al. [6*] | 2021 | South Africa | Prospective | 188 | 117 | 71 | Pediatric surgery |
| 7 | Shuaibu IY et al. [7*] | 2021 | Nigeria | Retrospective | 99 | 52 | 47 | Head and Neck surgery |
| 8 | Reynolds RA et al. [8*] | 2021 | Zambia | Retrospective | 75 | 41 | 34 | Neurosurgery |
| 9 | Purcell LN et al. [9*] | 2021 | Malawi | Retrospective | 1904 | 1065 | 839 | Burn |
| 10 | Khirallah MG et al. [10*] | 2021 | Egypt | Prospective | 75 | 47 | 28 | Pediatric surgery |
| 11 | Badejo OA et al. [11*] | 2021 | Nigeria | Retrospective | 12 | 5 | 7 | Neurosurgery |
| 12 | Allorto N et al. [12*] | 2021 | South Africa | Retrospective | 620 | 344 | 276 | Burn |
| 13 | Yehya A et al. [13*] | 2020 | Egypt | Prospective | 64 | 40 | 24 | Pediatric surgery |
| 14 | Pindyck T et al. [14*] | 2020 | Sub-Saharan Africa | Prospective | 1017 | 603 | 414 | Pediatric surgery |
| 15 | Peiffer S et al. [15*] | 2020 | Ghana | Retrospective | 468 | 422 | 46 | Pediatric surgery |
| 16 | Harunani S et al. [16*] | 2020 | Kenya | Prospective | 140 | 95 | 45 | Pediatric surgery |
| 17 | Ammer S et al. [17*] | 2020 | Tunisia | Retrospective | 182 | 126 | 56 | Pediatric surgery |
| 18 | Akello W et al. [18*] | 2020 | Uganda | Prospective | 40 | 24 | 16 | Pediatric surgery |
| 19 | Abdul-Mumin A et al. [19*] | 2020 | Ghana | Retrospective | 347 | 177 | 165 | Pediatric surgery |
| 20 | Punchak et al. [20*] | 2019 | Uganda | Retrospective | 100 | 61 | 39 | Neurosurgery |
| 21 | Nilsson E et al. [21*] | 2019 | Ghana | Retrospective | 133 | 96 | 37 | Pediatric surgery |
| 22 | Maine RG et al. [22*] | 2019 | Malawi | Prospective | 764 | 609 | 155 | Pediatric surgery |
| 23 | Leidinger A et al. [23*] | 2019 | Zanzibar | Prospective | 19 | 10 | 9 | Neurosurgery |
| 24 | El Sayed Y et al. [24*] | 2019 | Egypt | Retrospective | 303 | 288 | 15 | Ophthalmology |
| 25 | Shalaby R et al. [25*] | 2018 | Egypt | Retrospective | 1284 | 918 | 366 | Pediatric surgery |
| 26 | Rusingiza et al. [26*] | 2018 | Rwanda | Retrospective | 8 | 3 | 5 | Cardiac Surgery |
| 27 | Negussie T et al. [27*] | 2018 | Ethiopia | Retrospective | 53 | 29 | 24 | Pediatric surgery |
| 28 | Hussein AA et al. [28*] | 2018 | Egypt | Retrospective | 123 | 91 | 32 | Renal transplant |
| 29 | ElSheemy MS et al. [29*] | 2018 | Egypt | Retrospective | 108 | 73 | 35 | Renal Transplant |
| 30 | Anyanwu LJ et al. [30*] | 2018 | Nigeria | Retrospective | 129 | 78 | 51 | Pediatric surgery |
| 31 | Lin BM et al. [31*] | 2017 | Democratic Republic of the Congo | Prospective | 1207 | 725 | 482 | Pediatric surgery |
| 32 | Accorsi S et al. [32*] | 2017 | Ethiopia | Retrospective | 105025 | 40031 | 64994 | Pediatric surgery |
| 33 | Swart ED et al. [33*] | 2017 | South Africa | Retrospective | 856 | 531 | 325 | Pediatric surgery |
| 34 | Badawy H et al. [34*] | 2017 | Egypt | Prospective | 15 | 10 | 5 | Pediatric surgery |
| 35 | Gilbert CE et al. [35*] | 2016 | Multiple | Retrospective | 547 | 322 | 225 | Ophthalmology |
| 36 | Conway JC et al. [36*] | 2015 | Multiple | Retrospective | 27880 | 16454 | 11426 | Liver transplant |
| 37 | Shalaby R et al. [37*] | 2015 | Egypt | Retrospective | 38 | 34 | 4 | Pediatric surgery |
| 38 | Mahdi BD et al. [38*] | 2015 | Tunisia | Prospective | 90 | 75 | 15 | Pediatric surgery |
| 39 | Atwa ZT et al. [39*] | 2014 | Egypt | Cross-sectional | 312 | 171 | 141 | Cardiac surgery |
| 40 | Talabi AO et al. [40*] | 2014 | Nigeria | Retrospective | 45 | 26 | 19 | Pediatric surgery |
| 41 | Ibrahim MM et al. [41*] | 2014 | Egypt | Prospective | 14 | 10 | 4 | Pediatric surgery |
| 42 | Ezomike UO et al. [42*] | 2014 | Nigeria | Retrospective | 22 | 15 | 7 | Pediatric surgery |
| 43 | Simon R et al. [43*] | 2013 | Tanzania | Cross-sectional | 150 | 104 | 46 | Trauma |
| 44 | Ademuyiwa AO et al. [44*] | 2012 | Nigeria | Retrospective | 129 | 104 | 25 | Pediatric surgery |
| 45 | Elsayed H et al. [45*] | 2012 | Egypt | Prospective | 200 | 131 | 69 | Renal Transplant |
| 46 | Shalaby R et al. [46*] | 2010 | Egypt | Retrospective | 52 | 35 | 17 | Pediatric surgery |
| 47 | Ameh EA et al. [47*] | 2009 | Nigeria | Prospective | 76 | 40 | 36 | Pediatric surgery |
| 48 | Helmy TE et al. [48*] | 2009 | Egypt | Retrospective | 18 | 14 | 4 | Pediatric surgery |
| 49 | Abantanga FA et al. [49*] | 2009 | Ghana | Prospective | 955 | 568 | 387 | Pediatric surgery |
| 50 | Odebode TO et al. [50*] | 2006 | Nigeria | Retrospective | 60 | 28 | 32 | Neurosurgery |
| 51 | Kaabachi O et al. [51*] | 2005 | Tunisia | Retrospective | 458 | 282 | 176 | Orthopedics |
| 52 | Abantanga FA and Wiafe-Addai [52*] | 1998 | Ghana | Prospective | 121 | 76 | 45 | Pediatric surgery |
| 53 | Abantanga FA [53*] | 1997 | Ghana | Prospective | 66 | 41 | 25 | Pediatric surgery |
| 54 | Badibanga B et al. [54*] | 1980 | Zaire | Case review | 26 | 19 | 7 | Pediatric surgery |

